# Quantifying donor-recipient mismatches using recipient-derived sources of donor DNA

**DOI:** 10.64898/2026.06.01.26354606

**Authors:** Nallakkandi Rajeevan, Gabriel C. Barsotti, Ashwani Kumar, Zeguo Sun, Anand Reghuvaran, Irina Tikhonova, E M Tanvir, Niketa Sareen, Ashley Swan, Richard Formica, Caleigh Mandel-Brehm, Arundati Rao, Whitney Besse, Maureen Miller, Laurine Bow, Bony De Kumar, Madhav C. Menon

**Affiliations:** Department of Biomedical Informatics & Data Science, Yale School of Medicine, New Haven & West Haven, CT, USA; Section of Nephrology, Department of Internal Medicine, Yale School of Medicine, New Haven & West Haven, CT, USA; Yale Center for Genome Analysis, Yale School of Medicine, New Haven & West Haven, CT, USA; Department of Surgery, Yale School of Medicine, New Haven & West Haven, CT, USA; Department of Pathology, Yale School of Medicine, New Haven & West Haven, CT, USA; Versiti Blood Research institute, Milwaukee, WI 53202, United States

**Keywords:** Kidney transplantation, donor–recipient genetic mismatch, non-HLA incompatibility, urine cell-pellet DNA, whole-exome sequencing

## Abstract

Non-HLA donor-recipient (D-R) genetic mismatches contribute to kidney allograft injury and long-term graft loss, but their clinical use is limited by the unavailability of donor DNA after transplantation. We tested whether non-invasively obtained, recipient-derived samples could be used to infer donor genotype and D-R mismatches. Genomic DNA (g-DNA) of 11 unselected kidney transplant recipients and donors underwent whole-exome sequencing (100x). Additional customized probes were added for intronic coverage (300x) of 55 targeted non-HLA genes of reported clinical relevance. Variants identified from sequencing results were compared with plasma cell-free DNA (cfDNA), urine cell-pellet DNA (U-DNA) obtained from the same recipients. Genome-wide-, exonic-, or non-synonymous exonic- mismatches in transmembrane or secreted proteins, and mismatches within target genes were benchmarked using donor g-DNA to generate mismatch scores for each D-R pair. Within each of these genomic scales of mismatch, U-DNA identified D-R mismatches significantly better than the corresponding cfDNA (P<0.001 for each comparison). U-DNA also identified gene-level mismatches in the *LIMS1* gene, and correctly inferred established donor-origin risk alleles, including *SHROOM3 and APOL1*. Our findings demonstrate proof-of-concept that U-DNA in tandem with recipient genome, can non-invasively infer relevant non-HLA loci/mismatches circumventing the need for the donor’s genomic DNA.

## Introduction

In kidney transplantation (KTx), long-term allograft survival remains an unrealized goal^1, 2^. A role for anti-donor allo-immunity is implicated in all allograft loss (GL) from rejection, but also in histologic graft damage (fibrosis)^3, 4^. Immune injury is primarily triggered by recipient T-cells interacting with “mismatched” donor human leukocyte antigen (HLA) molecules via direct or indirect pathways of allorecognition. In donor-recipient pairs (D-R), non-HLA mismatches also trigger activation of non-self-reactions due to polymorphic donor peptides. Genome-wide studies, based on Single-nucleotide polymorphism (SNP) arrays or by whole exome sequencing (WES), have demonstrated that non-HLA D–R genetic mismatches influence graft loss through rejection^2, 5-7^ or graft fibrosis^8^. Besides non-synonymous exonic variants generating polymorphic donor proteins, mechanisms involving non-coding loci are also reported^9^. For instance, systematic scanning of genome-wide mismatches identified parsimonious gene-level mismatches that associated with GL, out of proportion from the remaining genome. The top-ranked gene locus *LIMS1*^9^ in this analysis, was independently validated^10^, and showed intronic haplotypes that act as cis-eQTLs, potentially modulating T-cell function and/or fibrogenesis in case of a “directional” *LIMS1* mismatch. Regardless of non-HLA mismatches, variants intrinsic to donors also program graft epithelial injury responses (reviewed in ^11^). Several such donor variants also associate with native kidney CKD^12-16^. Hence, <u>non-HLA mismatches have clear relevance to GL, and individual non-HLA loci/mismatches could allow the development of precision therapeutics if consistently mapped</u>.

However, donor genetic analyses are limited by logistics as donor genotype is usually unavailable when evaluating recipients. For instance, even in multiple NIAID cohorts (n∼1028 recipients) where researchers were instructed to collect donor-DNA ^17, 18^, we observed that donor-DNA for genotyping was available only in 31-68% cases. In clinical practice, transplanted donor organs are frequently imported (17 to 60%)^19^, and even with local donors, standardized recommendations for donor-DNA storage may not exist. Therefore, at the transplant center caring for recipients, obtaining non-HLA-variant or mismatch data from donor genotype remains markedly under-utilized in the current paradigm.

To circumvent this issue, we conceived and tested an assay utilizing either recipient urine cell pellet DNA (U-DNA) or the donor-derived cell-free DNA (dd-cfDNA) in recipient plasma as the source for donor genotyping. dd-cfDNA has already shown promise as a marker of immune injury in KTx^20 21^ and is in clinical utility ^22, 23 24^. However, existing commercial platforms use multiplex PCR (3-14K SNPs) and/or Next-generation sequencing (NGS) to differentiate recipient-from dd-cfDNA. The specific loci evaluated in each platform for D-R genotype inference are unclear. Based on sequential work investigating D-R variants ^8, 12, 25^, we generated a customized pipeline using WES augmented with custom intron-spanning gene-specific probes to target specific D-R mismatches of potential clinical significance. We report here that combining recipient genotype with this assay from recipient urine cell pellets highly correlated with mismatch scores and donor genotypes benchmarked using actual D-R g-DNA.

## Methods

### Study design

Cohort: From 11 kidney recipients at the Yale Transplant Clinic, recipient genomic DNA (g-DNA) was extracted from the buffy coat and cell-free DNA (cfDNA) from corresponding plasma samples at the time of allograft biopsy. Urine samples were obtained in parallel for U-DNA assessment. U-DNA and blood samples were collected prior to biopsy.

### Probe design

Hybrid capture probes were designed against targets regions using a proprietary design algorithm from twist Bioscience technologies. More details about probe designing are available in supplementary materials section.

### Urine collection and processing

Urine samples were obtained prior to kidney biopsy, case-7 was recollected during routine clinical visit due to the lack of identifiable cellpellet. and processed to get urine pellet for U-DNA isolation (see supplementary materials)^26^.

### Donor DNA and cfDNA sequencing

In brief, donor-DNA for each participant was obtained from the Yale transplant HLA bank, and cfDNA was isolated from plasma or serum, quantified, and prepared into indexed sequencing libraries using standard commercial kits. Libraries were pooled, target-captured with custom probes, quality-controlled by qRT-PCR and fragment analysis. Final libraries were sequenced on an Illumina NovaSeq X Plus using 100 bp paired-end reads. Sequencing data were processed using Illumina RTA for base calling and CASAVA for demultiplexing and primary analysis (details in supplementary materials).

### Analysis pipeline (For details, see supplementary materials.)

Briefly, after doing QC, libraries were sequenced using a human comprehensive exome panel supplemented with custom probes (Twist Biosciences) to achieve ∼100× exome and ∼300× targeted coverage, including intronic regions of selected non-HLA genes relevant to KTx. Reads were aligned to GRCh38 with BWA and processed using GATK v4 for duplicate marking, base quality recalibration, and variant calling with HaplotypeCaller to identify SNPs and INDELs.

### Annotation of SNP function

SNPs were annotated for genomic location using the UCSC known Gene transcriptome (Bioconductor v3.22) and for functional effects using Ensembl VEP (hg38) ^27^. Transmembrane and secreted genes were obtained from the AFTP and SEPDB databases, respectively ^28, 29^.

### Definition of D-R mismatch at different genomic scales

A mismatch was defined when the donor carried an allele absent in the recipient, defined as single or homozygous mismatches, which were collapsed into a single binary variable (any mismatch). Mismatches were annotated per SNP for each donor–recipient (D–R) pair. Gene-level mismatch scores were calculated by summing mismatches across SNPs within each gene. Scores were also aggregated across genomic scales, reported as raw sums.

### Statistical Analysis

All two group comparisons were performed using the Mann–Whitney U test; = p < 0.05.

### Study Approval

Recipients were recruited after informed consent under Yale IRB #200002603, which permitted collection of blood and urine specimens for genomic analyses.

## Results

### Cohort Description

In this proof-of-concept study, a total of 11 unselected kidney transplant recipients were recruited. The clinical characteristics of recipients and their corresponding donors are in Table 1. Ten out of 11 participants underwent allograft biopsy on the enrollment day, performed either for surveillance purposes (2 of 11 participants) or for clinical indication (8 of 11 participants). One scheduled surveillance biopsy could not be performed due to bowel interposition. Biopsy diagnoses are summarized in Table S1; only one participant demonstrated evidence of rejection at enrollment. All 11 recipients provided g-DNA and plasma cfDNA (Figure 1a). Donor g-DNA was not retrievable in case 7 resulting only 10 D-R pairs for g-DNA mismatch analyses. In addition, g-DNA, cfDNA and U-DNA were collected from a healthy non-transplanted volunteer as a technical control. Table-S2 lists concentrations and purity of collected U-DNA samples. Three out of 10 D-R pairs whose U-DNA did not pass QC were not used, resulting in 7 D-R pairs for U-DNA analysis.

**Table 1.**
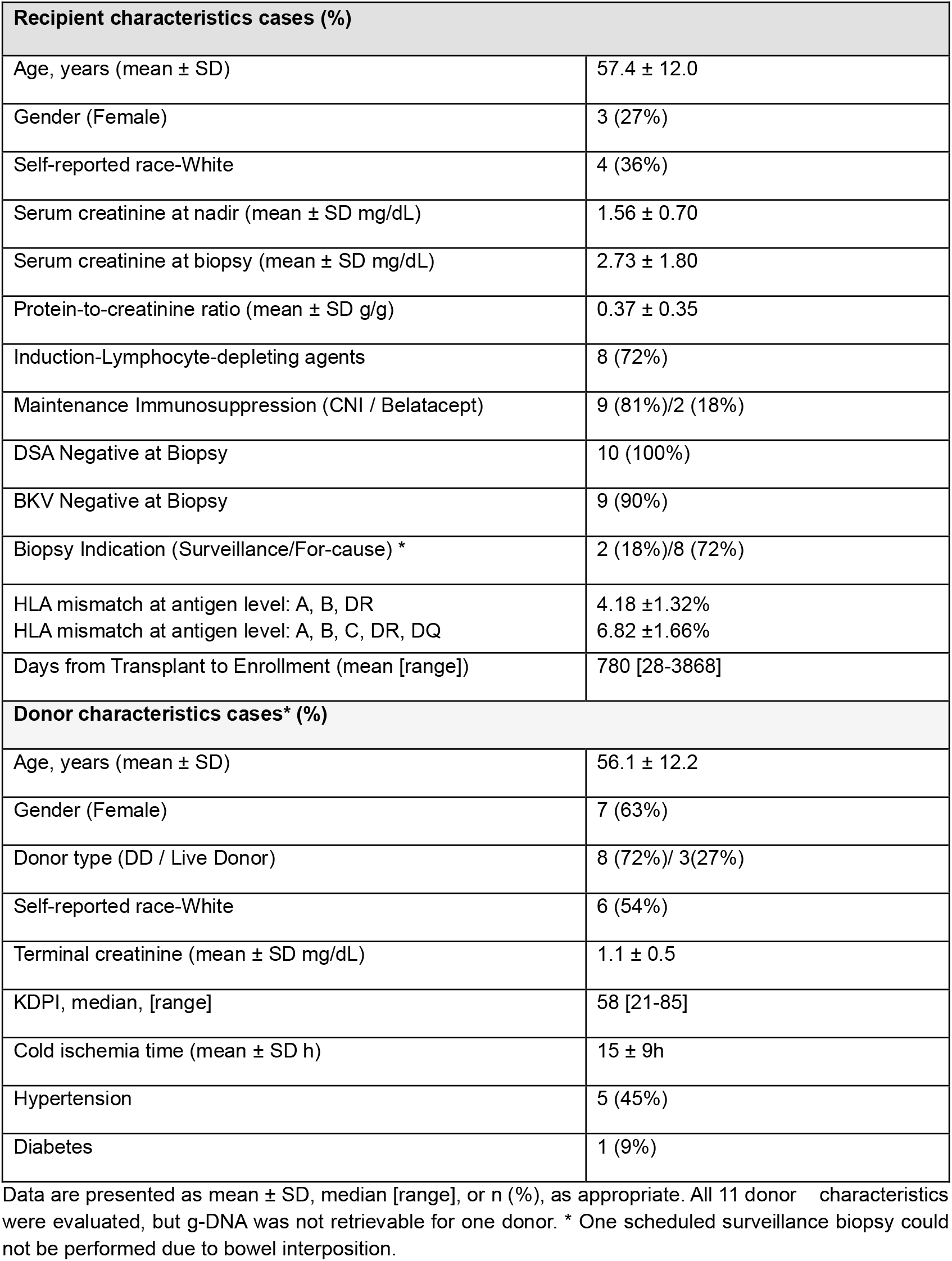
Recipient - Donor characteristics.

| **Recipient characteristics cases (%)** |  |
| --- | --- |
| Age, years (mean ± SD) | 57.4 ± 12.0 |
| Gender (Female) | 3 (27%) |
| Self-reported race-White | 4 (36%) |
| Serum creatinine at nadir (mean ± SD mg/dL) | 1.56 ± 0.70 |
| Serum creatinine at biopsy (mean ± SD mg/dL) | 2.73 ± 1.80 |
| Protein-to-creatinine ratio (mean ± SD g/g) | 0.37 ± 0.35 |
| Induction-Lymphocyte-depleting agents | 8 (72%) |
| Maintenance Immunosuppression (CNI / Belatacept) | 9 (81%)/2 (18%) |
| DSA Negative at Biopsy | 10 (100%) |
| BKV Negative at Biopsy | 9 (90%) |
| Biopsy Indication (Surveillance/For-cause) * | 2 (18%)/8 (72%) |
| HLA mismatch at antigen level: A, B, DR | 4.18 ± 1.32% |
| HLA mismatch at antigen level: A, B, C, DR, DQ | 6.82 ± 1.66% |
| Days from Transplant to Enrollment (mean [range]) | 780 [28-3868] |
| **Donor characteristics cases\* (%)** |  |
| Age, years (mean ± SD) | 56.1 ± 12.2 |
| Gender (Female) | 7 (63%) |
| Donor type (DD / Live Donor) | 8 (72%)/ 3(27%) |
| Self-reported race-White | 6 (54%) |
| Terminal creatinine (mean ± SD mg/dL) | 1.1 ± 0.5 |
| KDPI, median, [range] | 58 [21-85] |
| Cold ischemia time (mean ± SD h) | 15 ± 9h |
| Hypertension | 5 (45%) |
| Diabetes | 1 (9%) |

Data are presented as mean ± SD, median [range], or n (%), as appropriate. All 11 donor characteristics were evaluated, but g-DNA was not retrievable for one donor. \* One scheduled surveillance biopsy could not be performed due to bowel interposition.

**Figure 1:**
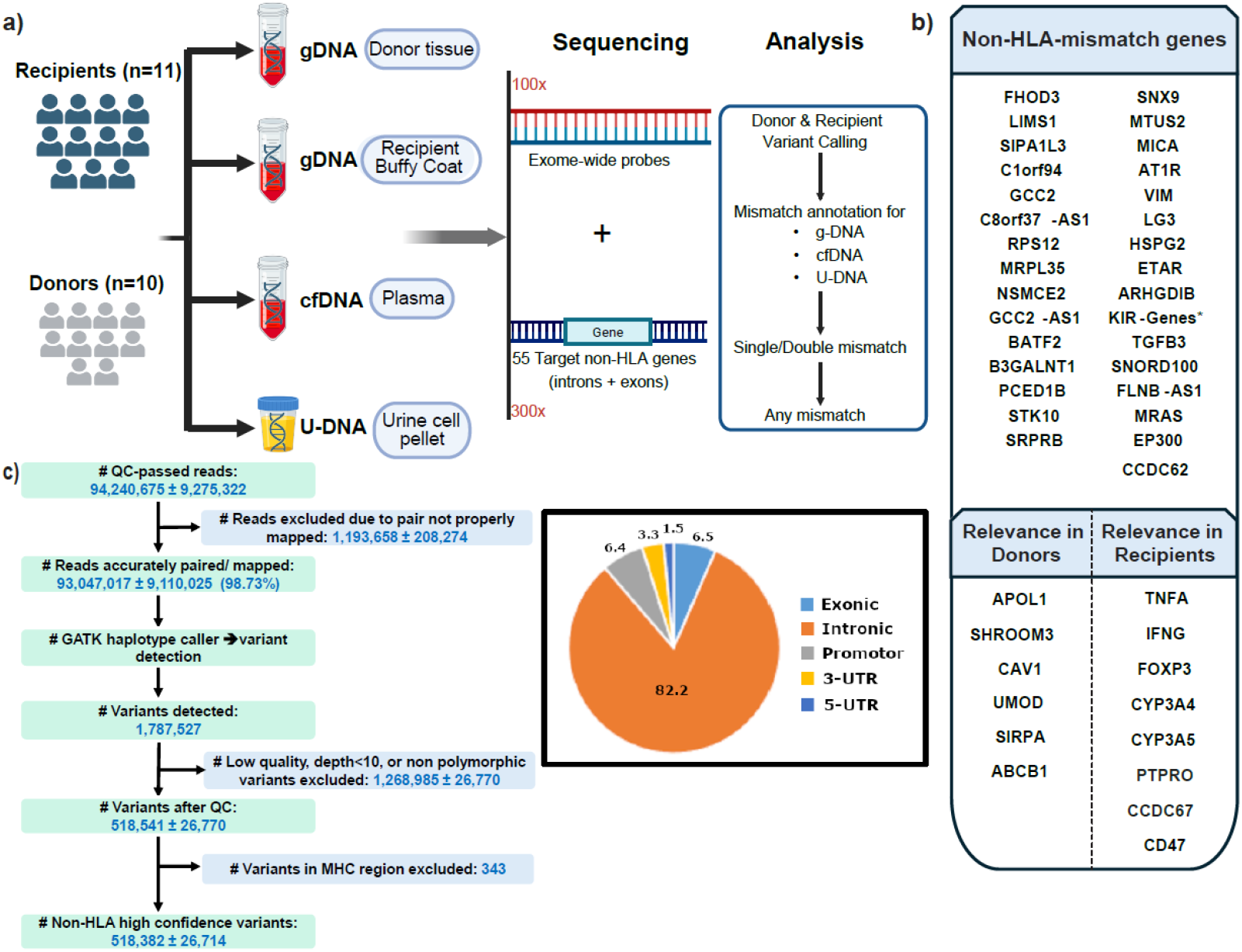
Study design, sequencing workflow, List of Non-HLA mismatched genes and QC. **(a)** Genomic DNA (g-DNA) was obtained from donor tissue and recipient buffy coat, while plasma cell-free DNA (cfDNA) and urinary cell pellet DNA (U-DNA) were isolated from recipient plasma and urine respectively (recipients n = 11; corresponding donors n=10*). Donor-recipient g-DNA, cfDNA/U-DNA underwent exome-wide sequencing (100x), across 55 targeted non-HLA genes. Variants were called in donors and recipients and annotated to identify donor-recipient mismatches across g-DNA, cfDNA, and U-DNA. *For one donor, there was no g-DNA available. **(b)** Selected non-HLA genes (N=45+10 KIR gene variants*) are grouped into relevant mismatch genes (variants in donors/recipients). These genes were selected based on published human transplant studies showing clinical association with KTx (Kidney transplant) outcomes. **(c)** Summary of read and variant quality control for a representative sample, including read mapping, variant calling, and exclusion of low-quality, non-informative, and MHC-region variants, resulting in high-confidence non-HLA variants. The pie chart shows the functional annotation distribution of retained variants.

### Genome-wide and target gene mismatch computation

Exome-wide sequencing in addition to sequencing selected non-HLA genes (n=55; Table S3, Figure 1b) using tiled probes to include introns was performed (see methods and Figure 1c for analytical pipeline). These 55 genes were selected based on published relevance to donors, recipients or as D-R mismatches^10, 12, 17^. Variant calling and mismatch annotations in g-DNA, cfDNA and U-DNA libraries are described in methods and in prior work ^17^.

For each D-R pair, mismatches were computed^17^ and benchmarked using donor- and recipient g-DNA. Briefly, the introduction of a new allele at a locus in a non-HLA gene where the recipient is homozygous was a “mismatch” (see methods). Using this approach, the total number of genome-wide mismatches *and* homozygous mismatches between the 10 D-R pairs are summarized in Table 2 [details in Supplementary excel]. An average of 73691 ± 13447 genome wide SNP mismatches (excluding HLA region) were identified from g-DNA in 10 D-R pairs [Figure 2a]. Interestingly, antigen level HLA mismatches between D-Rs using three- or five-antigens [Table-1], did not correlate with genome-wide non-HLA mismatches in our cohort [Figure S1]. As previously described ^5^, in each D-R pair, we also annotated and summarized *exonic*, and *non-synonymous exonic* SNP mismatches within secreted/ transmembrane proteins using UNIPROT^5^ [Table 2]. Finally, we tallied mismatches in the deeply sequenced 55 targeted gene regions which accounted for 2836 ±1166 mismatches [Table 2].

**Table 2.** Total mismatch and homozygous mismatches and their detection in g-DNA, cfDNA and U-DNA.

| Genomic Category | DNA Source | Total Mismatches (mean ± SD) | | Homo Mismatches (mean ± SD) | |
| --- | --- | --- | --- | --- | --- |
| | | #count ± SD | (mean% ± SD) | #count ± SD | (mean% ± SD) |
| Whole Genome (minus HLA) | g-DNA | 73691 ± 13447 | 100% | 15578±3720 | 100% |
| | cfDNA | 1375 ± 239 | 1.91 ± 0.39% | 19 ± 6.39 | 0.124 ± 0.044 % |
| | U-DNA | 51354 ± 23437 | 75.89 ± 35.12% | 650 ± 779 | 4.4 ± 5.28 % |
| Secreted/Transmembrane | g-DNA | 33272 ± 6052 | 100% | 7014 ± 1680 | 100% |
| | cfDNA | 577 ± 94 | 1.77 ± 0.34% | 8.3 ± 5.14 | 0.11 ± 0.07% |
| | U-DNA | 23458 ± 10685 | 76.87± 35.47% | 265 ± 329 | 4.0 ± 4.9% |
| Exonic non-synonymous<br>Secreted/Transmembrane | g-DNA | 9858 ± 1616 | 100% | 2086 ± 494 | 100% |
| | cfDNA | 117 ± 21 | 1.2 ± 0.30% | 1.3 ± 1.05 | 0.06 ± 0.05% |
| | U-DNA | 7121 ± 3272 | 77.93 ± 36.46% | 73 ± 80.70 | 3.10 ± 3.83% |
| Target genes | g-DNA | 2836 ± 1166 | 100% | 538 ± 272 | 100% |
| | cfDNA | 80 ± 30 | 2.89 ± 0.76% | 0.6± 0.7 | 0.13± 0.17 % |
| | U-DNA | 1787 ± 1228 | 76.50 ± 25.47% | 11 ± 13.8 | 2.91 ± 3.7 % |

Mean ± SD total and homozygous mismatch counts and percentages in g-DNA, cfDNA, and U-DNA across genomic categories; percentages are normalized to g-DNA (100%) within each category

**Figure 2:**
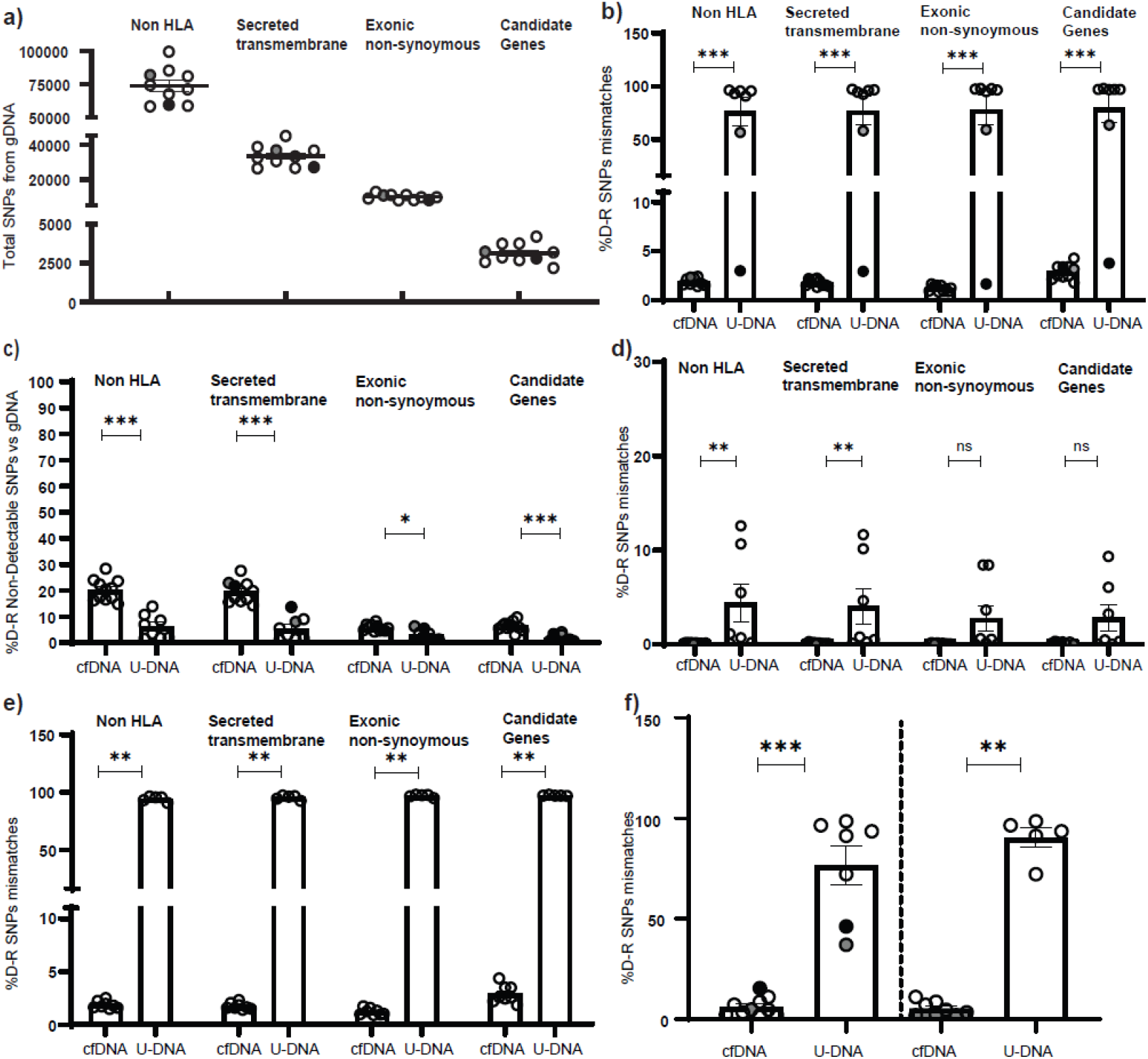
Distribution and detection of donor-recipient SNPs across gene categories in g-DNA, cfDNA, and U-DNA. **(a)** Total number of SNPs across gene categories in genomic DNA (g-DNA). **(b)** Percentage of donor–recipient (D–R) SNP mismatches detected in cfDNA and U-DNA across gene categories. **(c)** Percentage of SNPs not detected in cfDNA or U-DNA across gene categories. **(d)** Percentage of donor-recipient (D-R) homozygous SNP mismatches detected in cfDNA and U-DNA across gene categories **(e)** Percentage of donor–recipient (D-R) SNP mismatches detected in cfDNA and U-DNA across gene categories after exclusion of cases 2 and 10 (outliers). **(f)** Percentage of donor–recipient (D-R) SNP mismatches in the *LIMS1* gene, shown with outliers included (right) and excluded (left). *Outlier cases are indicated as follows: Case 2 is shown in gray, and Case 10 is shown as black dots. [All two group comparisons were performed using the Mann–Whitney U test; * = p < 0.05]

### Genome-wide mismatch inferences from cfDNA and U-DNA

Next, we aligned U-DNA and plasma cfDNA sequencing results from each recipient with the respective donor- and recipient-g-DNA sequencing results. At each position, SNPs aligned with recipient- or donor-g-DNA were first summarized. SNPs that were *not* aligned with recipient, but included an allele aligned with donor g-DNA in each U-DNA and cfDNA sample represented true D-R mismatches [summarized in Table 2, Figure 2b and tabulated for each D-R pair in Supplementary excel sheet]. In this manner, U-DNA identified 75.89% ± 35.12 of mismatches. On the other hand, cfDNA predominantly correlated with recipient allele-type and was able to represent 1.91% ± 0.39 of g-DNA-benchmarked D-R mismatches.

We then quantified SNPs that were identifiable in recipient g-DNA, but not in the donor g-DNA, and were also undetectable in the corresponding U-DNA and/or cfDNA. To understand baseline levels of such non-detection of recipient g-DNA SNPs in the same individual’s U-DNA and cfDNA, we utilized a non-transplanted control. We identified non-detection rates 5.94% to 10.73% across genome-wide, non-synonymous and targeted gene mismatches in the control U-DNA and cfDNA [Figure S2]. Among our D-R pairs, we observed that the non-detection rate of recipient g-DNA-identified SNPs ranged from 3.62% (1.48 to 5.43%) in U-DNA and 10.16% (5.61 to 19.77%) in cfDNA [Figure 2c].

U-DNA also significantly better represented benchmarked D-R mismatches in exonic-, or non-synonymous exonic-SNPs within secreted/ transmembrane proteins, as well as within the 55 targeted genes, vs cfDNA [Figure 2b]. Taken together, U-DNA sequences that were misaligned with recipient-g-DNA were highly representative of true D-R mismatches, while cfDNA was less reliable for mismatch inference. Since both U-DNA and cfDNA represent a mixture of donor- and recipient DNA, homozygous mismatches were poorly identified in both U-DNA and cfDNA [Figure 2d].

### Outlier cases in U-DNA mismatch results

As shown in Figure 2b, two out of seven cases with U-DNA showed lower representation of donor-sequences to infer D-R mismatch.

#### Case-2

[marked in Figure 2 as solid grey dot], had leukocyturia (4+ on dipstick; >30 WBC/high power field) with a positive urine culture for multidrug-resistant E. coli. We reasoned that greater proportion of leukocytes in this urine pellet – which are recipient derived – reduced the donor representation of the genome in U-DNA.

#### Case-10

[marked in Figure 2 as solid black dot] had undergone a combined liver–kidney transplant for acute hepatorenal syndrome. Post-transplantation, native kidney recovery in this patient was clinically suggested by remarkably low serum creatinine levels (<0.8mg/dL). In agreement, native kidney ultrasound showed normal sized kidneys with normal echotexture four years after transplantation. The contribution of recipient cells from the native kidneys to urine pellet was likely substantial in this patient, again reducing the U-DNA representation of donor genome.

Hence, these two outlier cases with inconsistent U-DNA results could ideally be identified *a priori* from clinical history or urine microscopy. Without these two outliers, U-DNA showed higher sensitivity in identifying genome-wide- or targeted gene-mismatches of 94.32%± 2.30 [Figure 2e and Table S4].

### Donor Recipient mismatch calculation in the absence of donor g-DNA

As mentioned, Case-7 had a transplant 13 years ago, and the corresponding donor g-DNA was not retrievable, exemplifying the limitation of donor-DNA even within the curated Yale biobank. Donor g-DNA based genotype for benchmarking D-R mismatches was thus unavailable in Case-7.We tested if U-DNA could reveal mismatch information in this recipient. The number of D-R mismatches identified using U-DNA in Case-7 was comparable to genome-wide- or targeted D-R mismatches observed from g-DNA and U-DNA in other cases (where donor g-DNA was available), while cfDNA detected lower numbers of mismatches [Figure S3], further supporting the utility of U-DNA for mismatch assessment.

### Inference of selected gene non-HLA D-R mismatches and donor-allele types using U-DNA

We next focused on the inference of selected variants that have shown relevance when present in the donor-, or as D-R mismatches, in both cases requiring ascertainment of donor genotype.

#### LIMS1

Both *gene-wide* LIMS1 mismatches, and mismatches at an intronic SNP [rs893403 with a “directional’ risk mismatch = A-allele donor to G/G recipient, i.e. minor A-allele introduced by donor into G/G recipients], have associated with rejection ^9, 30^, and/or GL^17^. U-DNA results correctly identified 2 mismatched D-Rs and reported Case-1 as a high-risk *LIMS1*-mismatch [Table 3]. Interestingly, in Case-7 where donor g-DNA was not available for benchmarking, U-DNA predicted the existence of a high risk LIMS1-D-R mismatch at this locus. Gene-wide LIMS1 mismatches (incorporating all identifiable SNPs within gene bounds) were also significantly better identified by U-DNA than cfDNA, with or without outlier cases [Figure 2f].

**Table 3.** Donor–recipient mismatch status at rs893403 *(LIMS1–GCC2*) determined using donor g-DNA, cfDNA, and U-DNA.

| D-R pair | rs893403<br>(*LIMS1–GCC2*) |  | Mismatch<br>(Y/N) | Risk mismatch<br>(Y/N) | cfDNA<br>genotype | U-DNA<br>genotype | Mismatch<br>from U-DNA<br>(Y/N) | Risk mismatch<br>from Urine (Y/N) |
| --- | --- | --- | --- | --- | --- | --- | --- | --- |
|  | Donor<br>genotype | Recipient<br>genotype |  |  |  |  |  |  |
| Case-1 | G/A | G/G | Y | Y | G/G | G/A | Y | Y |
| Case-2 | A/A | G/A | N | N | G/A | G/A | N | N |
| Case-4 | G/A | A/A | Y | N | A/A | G/A | Y | N |
| Case-7* | - | G/G | - | - | G/G | G/A | Y | Y |
| Case-8 | A/A | G/A | N | N | G/A | G/A | N | N |
| Case-9 | G/G | G/A | N | N | G/A | G/A | N | N |
| Case-10 | A/A | G/A | N | N | G/A | G/A | N | N |
| Case-11 | A/A | G/A | N | N | G/A | G/A | N | N |

Cases- 3, 5, and 6 were excluded due to insufficient urine DNA quality; \*Case- 7 lacked donor DNA. Y- Yes and N- No

#### SHROOM3

[rs17319721; risk allele= A-allele (Ref=G-allele)]. This intronic variant has been repeatedly associated with CKD^15, 16^, and donor risk genotype has been linked with allograft fibrosis and reduced GFR^12, 31^. We used U-DNA to check the inference of donor genotype comparing with donor-g-DNA [see Table S5]. In this pilot cohort, no D-R pair existed where donor g-DNA had the risk A-allele at this locus, where the corresponding recipient did not. However, U-DNA correctly inferred new alleles at this locus which did not align with recipient g-DNA, and aligned with donor g-DNA (cases 1 and 11).

#### APOL1 G2

[G2= rs71785313, a 6bp deletion]. One recipient- and a second D-R pair-(both donor and recipient) had G2 allele heterozygosity identifiable from their g-DNA. U-DNA identified the presence of this G2-allele as a heterozygote in the urine in both instances. We did not have any D-R pairs in this cohort where donor alone had G2 genotype [Table S6].

## Discussion

While D-R genetic studies have shown that non-HLA mismatches influence rejection and long-term allograft loss ^11^, obtaining donor-DNA at the recipient’s bedside for risk estimation after transplantation represents a hurdle. A significant proportion (∼55%) of deceased-donor transplants in the US result from imported organs, which has increased under the new KAS250 allocation system^19, 32^. Obtaining donor DNA for imported organs is thus particularly challenging, although even locally procured donors, or live donors do not have uniform policies for donor-DNA biobanking. Even in well-designed NIH-funded cohort studies where donor-DNA-collection was expected by protocol, donor DNA was obtained only from 31% to 68% ^12, 17, 18^. While paraffin embedded allograft biopsy is a potential source for donor genotyping, this involves tissue exhaustion, and generates fragmented libraries requiring dedicated sequencing approaches ^33-35^. Therefore, we evaluated and compared two non-invasive sources of donor-DNA sequences: recipient cfDNA and U-DNA to infer donor genotype and quantify D-R mismatches without directly requiring donor g-DNA. We surmised that circumventing this limitation would facilitate clinical translation of D-R mismatch assessments that already have clear relevance to outcomes.

We performed whole-exome sequencing with augmented targeted sequencing for a priori selected genes and adapted a pipeline for variant calling, comparing sequences obtained from U-DNA/cfDNA with the background donor- and recipient g-DNA sequences. We report that U-DNA identified donor-variants and D-R mismatch patterns significantly better than cfDNA, and approximating g-DNA. Contrarily, cfDNA was nearly entirely representative of the recipient genome, with higher noise-to-signal ratios. Although graft injury ^36^ increases the proportion of donor-origin cfDNA, dd-cfDNA is overall present at low concentrations, is fragmented into short molecules, and is susceptible to chemical and enzymatic degradation^37^. Our data also suggest that these fragmented cfDNA may randomly lack donor sequences from some target genes where mismatches have established clinical connotation. In contrast, U-DNA from urine cell-pellets-a mixture of kidney-resident cells (donor origin), as well as inflammatory/bladder cells (recipient), consistently allowed estimating non-HLA mismatches within selected gene regions. We also tested non-synonymous-SNPs in transmembrane/secreted proteins given their plausible clinical role^5^. Importantly, U-DNA also accurately inferred known risk loci in Shroom3, Lims1-Gcc2 and Apol1-G2. While Apol1-G1 risk alleles are also likely to be correctly inferred in U-DNA, false positive results occurred in our study due to a BAC-transgenic mouse model co-incidentally used in our lab (data not shown). We believe that this temporary error is unique and thus non-transferable to another laboratory. Overall, we provide rationale for developing U-DNA assays for D-R mismatch inference in the recipient, as well as identifying specific donor variants involved in allograft disease.

We note that we used deep sequencing for proof-of-concept here, subsequent assays could use SNP-genotyping or SNP-PCR platforms which are relatively inexpensive. Targeted multiplex PCR platforms will not require bioinformatic expertise and may be feasible for low resource settings. Further validation in larger cohorts, along with refinement of analytical pipelines will better establish diagnostic sensitivity, specificity and reproducibility. Although allograft biopsy extracted DNA could provide donor genotype, U-DNA that can be obtained on any recipient visit, is a non-invasive alternative, and could bridge a critical gap in clinical application non-HLA mismatch data. Our work also identified limitations inherent to this approach. First, leukocyturia can dilute down the donor-DNA content in U-DNA and reduce reliable mismatch calls. Microbial contamination can introduce unrelated DNA sequences yielding false-positive alignments at loci with homologous regions in microbial genomes. Hence, urine samples with significant leukocyturia or bacteriuria in urinalyses could be avoided for U-DNA estimation of mismatch. Second, in multi-organ transplant recipients with residual native kidney function, U-DNA may predominantly originate from the native rather than the transplanted kidney, as seen in our combined liver-kidney transplant case. Finally, we used a targeted gene list based on prior non-HLA work, but the U-DNA platform allows expansion to include any variants of significance.

In conclusion, we report the utility of U-DNA in tandem with recipient DNA for reliable identification of D-R mismatches at genome-wide, parsimonious- or gene-specific-scales providing proof-of-concept that donor genotype can be inferred non-invasively in the absence of banked donor-DNA, accelerating the translation of non-HLA donor-recipient mismatch information to the bedside.

## Data Availability

All data produced in the present study are available upon reasonable request to the authors

## Abbreviations

APOL1: Apolipoprotein L1
cfDNA: Cell-free DNA
CKD: Chronic kidney disease
dd-cfDNA: Donor-derived cell-free DNA
D-R: Donor–recipient
g-DNA: Genomic DNA
GFR: Glomerular filtration rate
GL: Graft loss
Gw: Genome-wide
HLA: Human leukocyte antigen
IF/TA: Interstitial fibrosis and tubular atrophy
KAS: Kidney Allocation System
KTx: Kidney transplantation
mH: Minor histocompatibility
NGS: Next-generation sequencing
QC: Quality control
SNP: Single nucleotide polymorphism
U-DNA: Urine cell-pellet DNA
WES: Whole-exome sequencing

## Data Availability Statement

The authors declare that all other relevant data supporting the findings of this study are available in this article and its Supplementary Information files. Additional data requests can be sent to Dr. Madhav Menon, the corresponding author.

## References

1. Hart A, Lentine KL, Smith JM, et al. OPTN/SRTR 2019 Annual Data Report: Kidney. Am J Transplant. Feb 2021;21 Suppl 2:21–137. doi:10.1111/ajt.16502

2. Arthur VL, Guan W, Loza BL, Keating B, Chen J. Joint testing of donor and recipient genetic matching scores and recipient genotype has robust power for finding genes associated with transplant outcomes. Genet Epidemiol. Nov 2020;44(8):893–907. doi:10.1002/gepi.22349

3. Modena BD, Kurian SM, Gaber LW, et al. Gene Expression in Biopsies of Acute Rejection and Interstitial Fibrosis/Tubular Atrophy Reveals Highly Shared Mechanisms That Correlate With Worse Long-Term Outcomes. American journal of transplantation : official journal of the American Society of Transplantation and the American Society of Transplant Surgeons. Jul 2016;16(7):1982–98. doi:10.1111/ajt.13728

4. El-Zoghby ZM, Stegall MD, Lager DJ, et al. Identifying specific causes of kidney allograft loss. Am J Transplant. Mar 2009;9(3):527–35. doi:10.1111/j.1600-6143.2008.02519.x

5. Reindl-Schwaighofer R, Heinzel A, Kainz A, et al. Contribution of non-HLA incompatibility between donor and recipient to kidney allograft survival: genome-wide analysis in a prospective cohort. Lancet. Mar 2 2019;393(10174):910–917. doi:10.1016/S0140-6736(18)32473-5

6. Pineda S, Sigdel TK, Chen J, Jackson AM, Sirota M, Sarwal MM. Novel Non-Histocompatibility Antigen Mismatched Variants Improve the Ability to Predict Antibody-Mediated Rejection Risk in Kidney Transplant. Front Immunol. 2017;8:1687. doi:10.3389/fimmu.2017.01687

7. Mesnard L, Muthukumar T, Burbach M, et al. Exome Sequencing and Prediction of Long-Term Kidney Allograft Function. PLoS Comput Biol. Sep 2016;12(9):e1005088. doi:10.1371/journal.pcbi.1005088

8. Zhang Z, Menon MC, Zhang W, et al. Genome-wide non-HLA donor-recipient genetic differences influence renal allograft survival via early allograft fibrosis. Kidney international. Sep 2020;98(3):758–768. doi:10.1016/j.kint.2020.04.039

9. Steers NJ, Li Y, Drace Z, et al. Genomic Mismatch at LIMS1 Locus and Kidney Allograft Rejection. N Engl J Med. May 16 2019;380(20):1918–1928. doi:10.1056/NEJMoa1803731

10. Kiryluk K, Steers NJ, Gharavi AG. Genomic Mismatch at LIMS1 Locus and Kidney Allograft Rejection. Reply. N Engl J Med. Aug 29 2019;381(9):e16. doi:10.1056/NEJMc1908072

11. Jethwani P, Rao A, Bow L, Menon MC. Donor-Recipient Non-HLA Variants, Mismatches and Renal Allograft Outcomes: Evolving Paradigms. Front Immunol. 2022;13:822353. doi:10.3389/fimmu.2022.822353

12. Menon MC, Chuang PY, Li Z, et al. Intronic locus determines SHROOM3 expression and potentiates renal allograft fibrosis. J Clin Invest. Jan 2015;125(1):208–21. doi:10.1172/JCI76902

13. Genovese G, Friedman DJ, Ross MD, et al. Association of trypanolytic ApoL1 variants with kidney disease in African Americans. Science. Aug 13 2010;329(5993):841–5. doi:10.1126/science.1193032

14. Palanisamy A, Reeves-Daniel AM, Freedman BI. The impact of APOL1, CAV1, and ABCB1 gene variants on outcomes in kidney transplantation: donor and recipient effects. Pediatr Nephrol. Sep 2014;29(9):1485–92. doi:10.1007/s00467-013-2531-7

15. Kottgen A, Pattaro C, Boger CA, et al. New loci associated with kidney function and chronic kidney disease. Nat Genet. May 2010;42(5):376–84. doi:10.1038/ng.568

16. Loeb GB, Kathail P, Shuai RW, et al. Variants in tubule epithelial regulatory elements mediate most heritable differences in human kidney function. Nat Genet. Oct 2024;56(10):2078–2092. doi:10.1038/s41588-024-01904-6

17. Sun Z, Zhang Z, Banu K, et al. Multiscale genetic architecture of donor-recipient differences reveals intronic LIMS1 mismatches associated with kidney transplant survival. J Clin Invest. Nov 1 2023;133(21)doi:10.1172/JCI170420

18. Hricik DE, Armstrong B, Alhamad T, et al. Infliximab Induction Lacks Efficacy and Increases BK Virus Infection in Deceased Donor Kidney Transplant Recipients: Results of the CTOT-19 Trial. J Am Soc Nephrol. Jan 1 2023;34(1):145–159. doi:10.1681/ASN.2022040454

19. Alhamad T, Marklin G, Ji M, Rothweiler R, Chang SH, Wellen J. One-Year Experience With the New Kidney Allocation Policy at a Single Center and an OPO in the Midwestern United States. Transpl Int. 2022;35:10798. doi:10.3389/ti.2022.10798

20. Halloran PF, Reeve J, Madill-Thomsen KS, et al. The Trifecta Study: Comparing Plasma Levels of Donor-derived Cell-Free DNA with the Molecular Phenotype of Kidney Transplant Biopsies. Journal of the American Society of Nephrology : JASN. Feb 2022;33(2):387–400. doi:10.1681/ASN.2021091191

21. Bloom RD, Bromberg JS, Poggio ED, et al. Cell-Free DNA and Active Rejection in Kidney Allografts. J Am Soc Nephrol. Jul 2017;28(7):2221–2232. doi:10.1681/ASN.2016091034

22. Agbor-Enoh S, Shah P, Tunc I, et al. Cell-Free DNA to Detect Heart Allograft Acute Rejection. Circulation. Mar 23 2021;143(12):1184–1197. doi:10.1161/CIRCULATIONAHA.120.049098

23. Levitsky J, Kandpal M, Guo K, Kleiboeker S, Sinha R, Abecassis M. Donor-derived cell-free DNA levels predict graft injury in liver transplant recipients. Am J Transplant. Feb 2022;22(2):532–540. doi:10.1111/ajt.16835

24. Bunnapradist S, Leca N, Zaky ZS, et al. Associations between donor-derived cell-free DNA dynamics and clinical outcomes after kidney allograft rejection: A prospective, multicenter study. Am J Transplant. Dec 2025;25(12):2543–2553. doi:10.1016/j.ajt.2025.07.2470

25. Zhang Z, Sun Z, Fu J, et al. Recipient APOL1 risk alleles associate with death-censored renal allograft survival and rejection episodes. J Clin Invest. Nov 15 2021;131(22)doi:10.1172/JCI146643

26. Barsotti GC, Luciano R, Kumar A, et al. Rationale and Design of a Phase 2, Double-blind, Placebo-Controlled, Randomized Trial Evaluating AMP Kinase-Activation by Metformin in Focal Segmental Glomerulosclerosis. Kidney Int Rep. May 2024;9(5):1354–1368. doi:10.1016/j.ekir.2024.02.006

27. McLaren W, Gil L, Hunt SE, et al. The Ensembl Variant Effect Predictor. Genome Biol. Jun 6 2016;17(1):122. doi:10.1186/s13059-016-0974-4

28. Pei J, Cong Q. AFTM: a database of transmembrane regions in the human proteome predicted by AlphaFold. Database (Oxford). Mar 14 2023;2023 doi:10.1093/database/baad008

29. Wang R, Ren C, Gao T, et al. SEPDB: a database of secreted proteins. Database (Oxford). Feb 12 2024;2024 doi:10.1093/database/baae007

30. Caliskan Y, Karahan G, Akgul SU, et al. LIMS1 risk genotype and T cell-mediated rejection in kidney transplant recipients. Nephrol Dial Transplant. Nov 9 2021;36(11):2120–2129. doi:10.1093/ndt/gfab168

31. Yan L, Li Y, Tang JT, et al. The influence of living donor SHROOM3 and ABCB1 genetic variants on renal function after kidney transplantation. Pharmacogenet Genomics. Jan 2017;27(1):19–26. doi:10.1097/FPC.0000000000000251

32. Puttarajappa CM, Hariharan S, Zhang X, et al. Early Effect of the Circular Model of Kidney Allocation in the United States. J Am Soc Nephrol. Jan 1 2023;34(1):26–39. doi:10.1681/ASN.2022040471

33. Cazzato G, Caporusso C, Arezzo F, et al. Formalin-Fixed and Paraffin-Embedded Samples for Next Generation Sequencing: Problems and Solutions. Genes (Basel). Sep 23 2021;12(10)doi:10.3390/genes12101472

34. Stiller M, Sucker A, Griewank K, et al. Single-strand DNA library preparation improves sequencing of formalin-fixed and paraffin-embedded (FFPE) cancer DNA. Oncotarget. Sep 13 2016;7(37):59115–59128. doi:10.18632/oncotarget.10827

35. Robbe P, Popitsch N, Knight SJL, et al. Clinical whole-genome sequencing from routine formalin-fixed, paraffin-embedded specimens: pilot study for the 100,000 Genomes Project. Genet Med. Oct 2018;20(10):1196–1205. doi:10.1038/gim.2017.241

36. Kueng N, Arcioni S, Sandberg F, et al. Comparison of methods for donor-derived cell-free DNA quantification in plasma and urine from solid organ transplant recipients. Front Genet. 2023;14:1089830. doi:10.3389/fgene.2023.1089830

37. Song P, Wu LR, Yan YH, et al. Limitations and opportunities of technologies for the analysis of cell-free DNA in cancer diagnostics. Nat Biomed Eng. Mar 2022;6(3):232–245. doi:10.1038/s41551-021-00837-3

