## Supplementary Materials for "Quantifying donor-recipient mismatches using recipient-derived sources of donor DNA"

**Appendix**


1. **Supplementary Methods –** pages 2-6
2. **Supplementary Tables –** pages 7-13
3. **Supplementary Figures –** pages 14-16
4. **Supplementary References –** pages 17-18

1. **Methods:**

**Study design:**

Cohort: In our pilot study, a total of 11 KTr from the Yale New Haven Hospital (YNHH) Transplant Clinic were recruited after informed consent under IRB protocol #200002603, which permitted enrollment as well as collection of blood and urine specimens for genomic analyses. Recipient genomic DNA (g-DNA) was extracted from recipient buffy coat. Corresponding plasma samples were used to extract cell-free DNA (cfDNA). During the time of clinically indicated allograft biopsy, peripheral blood was collected for cfDNA analysis, and urine samples were obtained in parallel for U-DNA assessment. U-DNA and cfDNA samples were collected prior to biopsy (Figure 1a).

**Probe design:**

Hybrid capture probes were designed against targets regions using a proprietary design algorithms from twist Bioscience technologies. Targeted probes were designed for 55 preselected gene. Probes were designed tiled to provide probe-probe overlap and to target both exonic and intronic regions, two probes are placed to avoid areas that will cause off target or low quality reads. The relative abundance of these probes was adjusted in design to provide uniform capture of the targets regardless of GC content. Probes were synthesized using Twist Bioscience's proprietary silicon-based technology. Each probe in the custom panel were assessed through NGS to correct GC boosting and probe sequence, a quality control step unique to the Twist Bioscience's NGS Target Enrichment Solutions.

**Urine collection and processing:**

Urine samples were obtained during clinically indicated transplant follow-up visits and samples were processed using a previously published workflow [1].

**Donor DNA and cfDNA sequencing:**

For each participant donor-DNA for donor genome genotyping was retrieved from the Yale transplant tissue typing laboratory. Sample Preparation: cfDNA from plasma or serum is isolated using the MagMax cfDNA Isolation Kit (Fisher, Part# A29319) according to the manufacturer's protocol. Extracted cfDNA samples are quantified and cfDNA purity were assessed via the cfDNA SceenTape assay (Agilent, Part# 5067-5630) on the TapeStation 4200 System (Agilent, Part#G2991AA). Samples with a yield of ≥1.0 ng/ul were used for further library preparation and sequencing. Library Preparation: 0.01μg of cfDNA was used to prepare sequencing libraries with Twist Library Preparation Kit (Twist Bioscience, Part# 104177) following the manufacturer's protocol. The adapter-ligated DNA fragments are PCR amplified using Twist UDI primers. During PCR, a unique 10-base index is inserted at both ends of each DNA fragment. Sample Target Capture: 1.5 μg of seven amplified, indexed libraries are pooled prior to capture. Pooled libraries were lyophilized, heat-denatured, and mixed with heat-denatured Twist Custom panel biotinylated DNA probes. Hybridizations are performed at 70°C for 16 hours. Once the capture was complete, the samples were mixed with streptavidin-coated beads and washed with a series of stringent buffers to remove non-specifically bound DNA fragments. The captured fragments are PCR amplified. Samples were quantified by qRT-PCR using a Kapa Library

Quantification Kit (Roche, Part #KK4854) and insert size distribution was determined with the Agilent Tape Station system. Samples with a yield of ≥0.5 ng/ul were used for sequencing. Flow Cell Preparation and Sequencing: Sample concentrations were normalized to 2nM and loaded onto an Illumina NovaSeq X Plus flow cell at a concentration that yields at least 40 M of passing filter clusters per sample. The loading concentration for libraries has been optimized to maximize both well occupancy and unique read output while limiting duplicates associated with patterned flow cell technology. Samples were sequenced using 100 bp paired-end sequencing reads according to Illumina protocols. The 10bp indexes were read during additional sequencing reads that automatically follow the completion of read 1. Data generated during sequencing runs were simultaneously transferred to the YCGA high-performance computing cluster. A positive control (prepared bacteriophage Phi X library) provided by Illumina was spiked into every lane at a concentration of 1% to monitor sequencing quality in real time.

**Data Analysis and Storage:** Signal intensities are converted to individual base calls during a run using the system's Real Time Analysis (RTA) software. Base calls were transferred from the machine's dedicated personal computer to the Yale High Performance Computing cluster via a 10 Gigabit network mount for downstream analysis. Primary analysis, including sample demultiplexing, is performed using Illumina's CASAVA 1.8.2 software suite. The data was returned to the user if the sample error rate was less than 2%. Data is retained on the cluster for at least 6 months, after which it is transferred to a tape backup system.

**Analysis pipeline:**

Library preparation was performed for each sample that passed QC. A total of 42 library preparations (39 samples from cases and 3 samples from a control) underwent sequencing at an average depth of 300X. Human comprehensive exome panel (Twist biosciences) were augmented with custom probes (designed and synthesized by Twist biosciences). Both panels were mixed in a ratio to get 100X coverage for general exomes while targeted regions were sequenced at a 300X coverage. These non-HLA genes were selected based on prior publications showing clinical relevance in transplant cohorts (Supplementary Table S3). Candidates were included to cover biologically plausible variants reported in donors, in recipients and previously described non-HLA mismatch variants. In these regions, probes were deliberately designed to cover intronic sequences within the bounds of these genes as several reported variants are intronic [2-4].

Read quality were assessed using FastQC [5] and adapter sequences were trimmed using BBDuck, a tool within the BBTools suite [6]. The quality-controlled reads were aligned to the homo sapiens assembly 38 reference genome (GRCh38) using Burrows-Wheeler Aligner (BWA-MEM version 0.7) [7]. For variant discovery, we followed the Best Practices Workflows detailed in genome analysis toolkit (GATK4) [8]. The BWA-aligned reads were pre-processed by first marking duplicates (MarkDuplicatesSpark) followed by computing and applying the base quality recalibration scores (BaseRecalibrator) in GATK. For variant calling, we used the GATK HaplotypeCaller in GVCF mode. This mode allowed us to find both SNPs and INDELs (Insertion/Deletions) simultaneously via a local de-novo assembly of haplotypes. This first created the GVCF files for each sample and then jointly genotyped for all samples, giving VCF files for each sample.

**Annotation of SNP function:**

SNPs were annotated in terms of genomic locations (exonic, intronic, promotor, etc.) using UCSC transcriptome database for known genes Bioconductor version 3.22 (TxDb.Hsapiens.UCSC.hg38. knownGene) and protein coding functions (synonymous, non-synonymous, frameshift, etc.) using Ensembl Variant Effect Predictor (VEP) for hg38 assembly [9]. The list of transmembrane genes were obtained from the AFTP (AlphaFold Transmembrane proteins) database [10] and the list of secreted genes were downloaded from the SEPDB database [11].

**Definition of D-R mismatch at different genomic scales:**

The D-R mismatch for each SNP was defined following the strategy as before A mismatch was defined as a donor carrying an allele that was not presented in the recipient. We consider mismatch derived from one “alien” allele introduced by donor as “single mismatch”, while two alleles introduced as “double mismatch”. The “single mismatch” and “double mismatch” were collapsed into a class named “any mismatch” (Figure 1). The mismatch score was calculated for each SNP among each donor-recipient pair. To define gene-level mismatch score, the mismatch status (0 for absence and 1 for presence) of all the SNPs within each annotated gene region were summed up. Similarly, to define mismatch score at different genomic scale, such as Gw, exonic, or all transmembrane or secreted gene regions, the mismatch status of all the SNPs within the corresponding genomic regions were summed as the raw score of the corresponding raw scores across D-R pairs.

**Supplementary Tables:**

**Table S1**

| **Kidney allograft biopsy findings (N = 10) *** | |
|---|---|
| **Biopsy finding** | **n (%)** |
| **Acute tubular injury (ATI)** | |
| Focal | 6 (60%) |
| Diffuse | 4 (40%) |
| Chronic calcineurin inhibitor toxicity | 2 (20%) |
| Polyomavirus nephropathy (Banff class 3) | 1 (10%) |
| Acute cellular rejection (Banff borderline) | 1 (10%) |
| **Glomerulosclerosis present** | 4 (40) |
| Percentage of glomerulosclerosis (> 20%) | 3 (30%) |
| **Interstitial fibrosis and tubular atrophy (IFTA)** | |
| Grade 2 or 3 | 3 (30%) |
| C4d positivity | 0 (0%) |

*One of 11 planned biopsies was not performed because of anatomical contraindication. IFTA graded per Banff criteria.

**Table S2**

| Tables S2. Participants U-DNA details | | | | | | | |
|---|---|---|---|---|---|---|---|
| **Study ID** | **Urine Vol (mL)** | **Con.(ng/ul)** | **260/280** | **260/230** | **Tape station (>1ng/ul)** | **DNA volume (ul)** | **Total DNA** |
| Case -1 | 100 | 19.40 | 1.84 | 1.7 | Yes | 25.00 | 485.00 |
| Case- 2 | 180 | 205.70 | 1.88 | 2.03 | Yes | 25.00 | 5142.50 |
| Case- 3 | 90 | 16.40 | 1.65 | 0.78 | **No** | 25.00 | 410.00 |
| Case- 4 | 60 | 181.70 | 1.85 | 2.18 | Yes | 25.00 | 4542.50 |
| Case- 5 | 180 | 2.50 | 1.22 | -21.84 | **No** | 25.00 | 62.50 |
| Case- 6 | 160 | 6.90 | 1.48 | 0.78 | **No** | 25.00 | 172.50 |
| Case- 7 | 60 | 4.80 | 1.47 | 0.71 | Yes | 25.00 | 120.00 |
| Case- 8 | 200 | 35.10 | 1.92 | 0.45 | Yes | 25.00 | 877.50 |
| Case- 9 | 70 | 12.60 | 1.64 | 1.82 | Yes | 25.00 | 315.00 |
| Case- 10 | 120 | 199.90 | 1.94 | 0.7 | Yes | 35.00 | 6996.50 |
| Case- 11 | 80 | 24.70 | 1.74 | 1.57 | Yes | 25.00 | 617.50 |

U-DNA yield and quality metrics: Urine volume, Con.(ng/ul)-DNA concentration, 260/280 -DNA purity, DNA volume - elution volume, and total DNA yield are shown for each study participant.

**Table S3**

| Non-HLA mismatch | | | |
|---|---|---|---|
| ***Gene*** | **Chromosome** | **Length (bp)** | **Key references** |
| ***FHOD3*** | 18 | 482,525 | [4] |
| ***LIMS1*** | 2 | 152,892 | [4,12] |
| ***SIPA1L3*** | 19 | 301,152 | [4] |
| ***C1orf94*** | 1 | 52,248 | [4] |
| ***GCC2*** | 2 | 60,278 | [4] |
| ***C8orf37-AS1*** | 8 | 541,308 | [4] |
| ***RPS12*** | 6 | 2,996 | [4] |
| ***MRPL35*** | 2 | 13,922 | [4] |
| ***NSMCE2*** | 8 | 275,303 | [4] |
| ***GCC2-AS1*** | 2 | 26,679 | [4] |
| ***BATF2*** | 11 | 9,101 | [4] |
| ***B3GALNT1*** | 3 | 21,490 | [4] |
| ***PCED1B*** | 12 | 157,061 | [4] |
| ***STK10*** | 5 | 146,273 | [4] |
| ***SRPRB*** | 3 | 37,460 | [4] |
| ***TGFB3*** | 14 | 24,951 | [4] |
| ***SNORD100*** | 6 | 76 | [4] |
| ***FLNB-AS1*** | 3 | 8,087 | [4] |
| ***MRAS*** | 3 | 57,888 | [4] |
| ***EP300*** | 22 | 87,468 | [13] |
| ***CCDC62*** | 12 | 52,872 | [4] |
| ***SNX9*** | 6 | 121,907 | [4] |
| ***MTUS2*** | 13 | 481,337 | [4] |
| ***MICA*** | 6 | 11,720 | [14] |
| ***AT1R*** | 3 | 55,000 | [15] |
| ***VIM*** | 10 | 17,228,241 | [16] |
| ***LG3*** | 14 | 21,321 | [17] |
| ***HSPG2*** | 1 | 115,067 | [18] |

| | | | |
|---|---|---|---|
| ***ETAR*** | 13 | 24,000 | [19] |
| ***ARHGDIB*** | 12 | 20,000 | [20] |
| ***KIR3DL2*** | 19 | 16,256 | [21] |
| ***KIR3DL1/S1*** | 19 | 14,311 | [21] |
| ***KIR2DS5*** | 19 | 15,021 | [21] |
| ***KIR2DS4*** | 19 | 15,700 | [21] |
| ***KIR2DS3*** | 19 | 14,405 | [21] |
| ***KIR2DS2*** | 19 | 14,335 | [21] |
| ***KIR2DS1*** | 19 | 14,015 | [21] |
| ***KIR2DL5B*** | 19 | 9,300 | [22] |
| ***KIR2DL5A*** | 19 | 9,400 | [22] |
| ***KIR2DL4*** | 19 | 10,000 | [23] |
| ***KIR2DL2/L3*** | 19 | 14,540 | [24] |
| ***KIR2DL1*** | 19 | 14,530 | [21] |
| **Donor gene variants** | | | |
| ***APOL1*** | 22 | 14,461 | [25] |
| ***SHROOM3*** | 4 | 10,891 | [2] |
| ***CAV1*** | 7 | 36,400 | [26] |
| ***UMOD*** | 16 | 20,000 | [27] |
| ***SIRPA*** | 20 | 48,000 | [28] |
| ***ABCB1*** | 7 | 120,000 | [29] |
| **Recipient gene variant** | | | |
| ***TNFA*** | 6 | 2,800 | [30] |
| ***IFNG*** | 12 | 5,000 | [31] |
| ***FOXP3*** | X | 49,250 | [32] |
| ***CYP3A4*** | 7 | 200,000 | [33] |
| ***CYP3A5*** | 7 | 200,000 | [33] |
| ***PTPRO*** | 12 | 280,000 | [34,35] |
| ***CCDC67*** | 11 | 93,329,971 | [35] |
| ***CD47*** | 3 | 48,771 | [36] |

The superscripted numbers are referred to in the reference list.

**Table S4**

| Total mismatch and homozygous mismatches and their detection in g-DNA, cfDNA and U-DNA | | | | | |
|---|---|---|---|---|---|
| **Genomic Category** | **DNA Source** | **Total mismatches (mean ± SD)** | | **Homozygous mismatches (mean ± SD)** | |
| | | **(mean count ± SD)** | **(mean% ± SD)** | **(mean count ± SD)** | **(mean% ± SD)** |
| Whole Genome (minus HLA) | g-DNA | 74450 ±13904 | 100% | 16232 ± 3828 | 100% |
| | cfDNA | 1304 ±160 | 1.79 ± 0.35% | 20 ± 5.16 | 0.134 ± 0.043% |
| | U-DNA | 62268 ± 7382 | 94.32 ± 2% | 902 ± 796 | 3.81 ± 5.16% |
| Secreted/Transmembrane | g-DNA | 33628 ± 6304 | 100% | 7346 ± 1717 | 100% |
| | cfDNA | 549 ± 64 | 1.67± 0.31% | 9.5 ± 4.89 | 0.13± 0.07% |
| | U-DNA | 28422 ± 3306 | 95.4 ± 1.9% | 367 ± 342 | 3.46 ± 4.85% |
| Exonic non-synonymous Secreted/Transmembrane | g-DNA | 9957 ± 1658 | 100% | 2185 ± 494 | 100% |
| | cfDNA | 111 ± 18 | 1.15 ± 0.30% | 1.12 ± 0.99 | 0.05 ± 0.04% |
| | U-DNA | 8657 ± 886 | 96.9 ± 1.2% | 87.2 ± 81.41 | 2.70 ± 3.72% |
| Target genes | g-DNA | 2792 ± 1313 | 100% | 531 ± 299 | 100% |
| | cfDNA | 75 ± 33 | 2.80 ± 0.82% | 0.6 ± 0.7 | 0.14 ± 0.18% |
| | U-DNA | 2625 ± 349 | 97.18 ± 0.3% | 16 ± 14 | 3.98 ± 3.93% |

The outliers' cases- 2 and 10 were excluded in this analysis. Mean ± SD total and homozygous mismatch counts and percentages in g-DNA, cfDNA, and U-DNA across genomic categories; percentages are normalized to g-DNA (100%) within each category

**Table S5**

| Genotype comparison for *SHROOM3* rs17319721 (risk allele A) across recipient donor g-DNA, cfDNA, and U-DNA | | | | |
|---|---|---|---|---|
| **rs17319721 (*SHROOM3*)** | **Recipient g-DNA** | **Donor g-DNA** | **cfDNA** | **U-DNA** |
| Case-1 | A/A | G/G | A/A | G/A |
| Case-2 | G/A | G/A | G/A | G/A |
| Case-4 | G/G | G/G | G/G | G/G |
| Case-7 * | G/A | - | G/A | G/G |
| Case-8 | G/G | G/G | G/G | G/G |
| Case-9 | G/A | G/A | G/A | G/A |
| Case-10 | G/A | G/G | G/A | G/A |
| Case-11 | A/A | G/A | A/A | G/A |

*No donor–recipient pair showed a donor risk A allele absent in the recipient; pair 7 lacked donor DNA

**Table S6*: APOL1* G2 (rs71785313) genotype calls**

| rs71785313 (***APOL1*-G2**) | **Recipient g-DNA** | **Donor g-DNA** | **cfDNA** | **U-DNA** |
|---|---|---|---|---|
| Case-3 | 0/1 | 0/1 | 0/1 | . |
| Case-11 | 0/1 | 0/0 | 0/1 | 0/1 |

Heterozygous genotypes (0/1) are shown for Cases 3 and 11; all other cases were homozygous reference (0/0). Deletion allele corresponds to 0/1 = ATAATTATAA/ATAA.

**Supplementary Figures:**

**Figure S1**

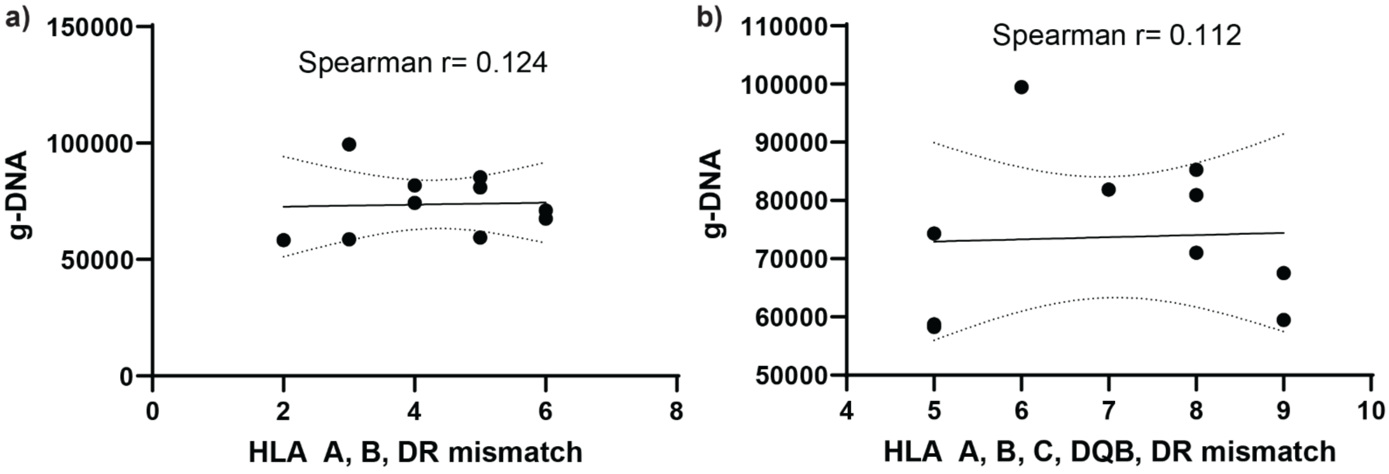


**Figure S1: Correlation plots showing antigen-level HLA mismatches between donor–recipient pairs using** (a) three antigens and (b) five antigens, in relation to genome-wide non-HLA mismatches in the present cohort.

**Figure S2**

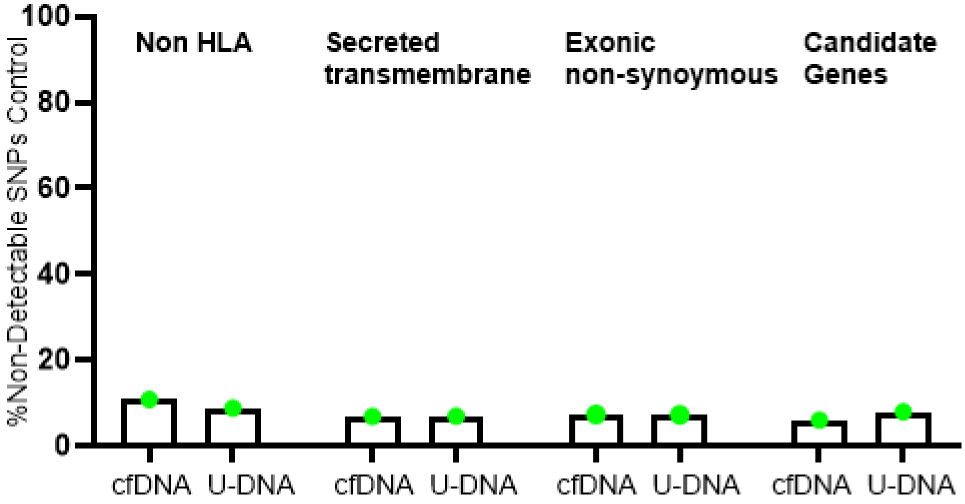


**Figure S2. Non-transplanted control sequencing data to assess non-detected rate.** Bar plot depicts percentage of g-DNA SNPs not detected in U-DNA and cfDNA across different gene annotation strategies in a control.

**Figure S3**

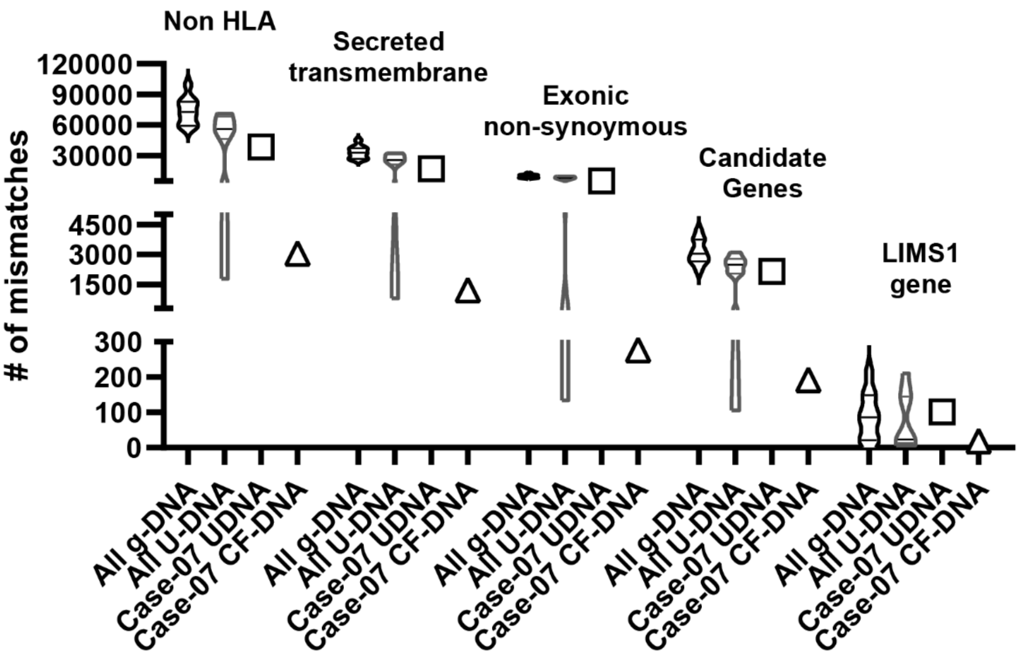


**Figure S3**: **Donor–recipient mismatch burden in Case-7 relative to other cases.** Violin plots showing donor–recipient (D-R) mismatches identified using genomic DNA (g-DNA) and urine-derived DNA (U-DNA) across all cases, with all distributions compared to Case-7. Mismatches are stratified by functional annotation (non-HLA variants, secreted/transmembrane genes, exonic non-synonymous variants, candidate genes, and *LIMS1*). For Case-7, mismatch counts derived from U-DNA (and cfDNA where available) are shown alongside global or mismatch estimates from g-DNA. Across all categories, Case-7 U-DNA–derived mismatch counts are comparable to those observed in other cases. All g-DNA-Violin plot; All U-DNA-Violin plot (truncated); Case 7 U-DNA-Square; Case 7 cfDNA-Triangle

**Reference List**


1. Barsotti GC, Luciano R, Kumar A, et al. Rationale and Design of a Phase 2, Double-blind, Placebo-Controlled, Randomized Trial Evaluating AMP Kinase-Activation by Metformin in Focal Segmental Glomerulosclerosis. Kidney Int Rep 2024;9(5):1354-1368. DOI: 10.1016/j.ekir.2024.02.006.
2. Menon MC, Chuang PY, Li Z, et al. Intronic locus determines SHROOM3 expression and potentiates renal allograft fibrosis. J Clin Invest 2015;125(1):208-21. DOI: 10.1172/JCI76902.
3. Kiryluk K, Steers NJ, Gharavi AG. Genomic Mismatch at LIMS1 Locus and Kidney Allograft Rejection. Reply. N Engl J Med 2019;381(9):e16. DOI: 10.1056/NEJMc1908072.
4. Sun Z, Zhang Z, Banu K, et al. Multiscale genetic architecture of donor-recipient differences reveals intronic LIMS1 mismatches associated with kidney transplant survival. J Clin Invest 2023;133(21). DOI: 10.1172/JCI170420.
5. Andrews S. FastQC: a quality control tool for high throughput sequence data. (http://www.bioinformatics.babraham.ac.uk/projects/fastqc ).
6. Grunden JMWaAM. Impact of BBDuk metagenomic read trimming and decontamination. . 2021. DOI: doi.org/10.25982/77705.1341/1779218.
7. Li H. Aligning sequence reads, clone sequences and assembly contigs with BWA-MEM. 2013. DOI: https://doi.org/10.48550/arXiv.1303.3997.
8. Van der Auwera GA, Carneiro MO, Hartl C, et al. From FastQ data to high confidence variant calls: the Genome Analysis Toolkit best practices pipeline. Curr Protoc Bioinformatics 2013;43(1110):11 10 1-11 10 33. DOI: 10.1002/0471250953.bi1110s43.
9. McLaren W, Gil L, Hunt SE, et al. The Ensembl Variant Effect Predictor. Genome Biol 2016;17(1):122. DOI: 10.1186/s13059-016-0974-4.
10. Pei J, Cong Q. AFTM: a database of transmembrane regions in the human proteome predicted by AlphaFold. Database (Oxford) 2023;2023. DOI: 10.1093/database/baad008.
11. Wang R, Ren C, Gao T, et al. SEPDB: a database of secreted proteins. Database (Oxford) 2024;2024. DOI: 10.1093/database/baae007.
12. Steers NJ, Li Y, Drace Z, et al. Genomic Mismatch at LIMS1 Locus and Kidney Allograft Rejection. N Engl J Med 2019;380(20):1918-1928. DOI: 10.1056/NEJMoa1803731.
13. Gong Y, Dou Y, Wang L, Wang X, Zhao Z. EP300 promotes renal tubular epithelial cell fibrosis by increasing HIF2alpha expression in diabetic nephropathy. Cell Signal 2022;98:110407. DOI: 10.1016/j.cellsig.2022.110407.
14. Risti M, Bicalho MD. MICA and NKG2D: Is There an Impact on Kidney Transplant Outcome? Front Immunol 2017;8:179. DOI: 10.3389/fimmu.2017.00179.
15. Abuzeineh M, Aala A, Alasfar S, Alachkar N. Angiotensin II receptor 1 antibodies associate with post-transplant focal segmental glomerulosclerosis and proteinuria. BMC Nephrol 2020;21(1):253. DOI: 10.1186/s12882-020-01910-w.
16. Latt KZ, Heymann J, Yoshida T, Kopp JB. Glomerular Kidney Diseases in the Single-Cell Era. Front Med (Lausanne) 2021;8:761996. DOI: 10.3389/fmed.2021.761996.
17. Surin B, Sachon E, Rougier JP, et al. LG3 fragment of endorepellin is a possible biomarker of severity in IgA nephropathy. Proteomics 2013;13(1):142-52. DOI: 10.1002/pmic.201200267.
18. Praditsap O, Ahsan NF, Nettuwakul C, et al. Whole exome sequencing reveals heparan sulfate proteoglycan 2 (HSPG2) as a potential causative gene for kidney stone disease in a Thai family. Urolithiasis 2024;53(1):7. DOI: 10.1007/s00240-024-01674-0.
19. Ma X, Liang Y, Chen W, Zheng L, Lin H, Zhou T. The role of endothelin receptor antagonists in kidney disease. Ren Fail 2025;47(1):2465810. DOI: 10.1080/0886022X.2025.2465810.
20. Senev A, Otten HG, Kamburova EG, et al. Antibodies Against ARHGDIB and ARHGDIB Gene Expression Associate With Kidney Allograft Outcome. Transplantation 2020;104(7):1462-1471. DOI: 10.1097/TP.0000000000003005.

21. Loga LI, Suharoschi R, Elec FI, et al. Orchestrating the Impact of KIR/HLA Interactions on Kidney Transplant. Int J Mol Sci 2024;25(15). DOI: 10.3390/ijms25158228.
22. Cisneros E, Moraru M, Gomez-Lozano N, Lopez-Botet M, Vilches C. KIR2DL5: An Orphan Inhibitory Receptor Displaying Complex Patterns of Polymorphism and Expression. Front Immunol 2012;3:289. DOI: 10.3389/fimmu.2012.00289.
23. Ding XF, Chen J, Ma HL, et al. KIR2DL4 promotes the proliferation of RCC cell associated with PI3K/Akt signaling activation. Life Sci 2022;293:120320. DOI: 10.1016/j.lfs.2022.120320.
24. Bao XJ, He J, Chen ZX, et al. [Study on the behavior of NK cell KIRs of donor/recipient pairs in HLA matched unrelated allo-HSCT]. Zhonghua Xue Ye Xue Za Zhi 2007;28(8):510-3. (https://www.ncbi.nlm.nih.gov/pubmed/18078124).
25. Zhang Z, Sun Z, Fu J, et al. Recipient APOL1 risk alleles associate with death-censored renal allograft survival and rejection episodes. J Clin Invest 2021;131(22). DOI: 10.1172/JCI146643.
26. Adam BA, Smith RN, Rosales IA, et al. Chronic Antibody-Mediated Rejection in Nonhuman Primate Renal Allografts: Validation of Human Histological and Molecular Phenotypes. Am J Transplant 2017;17(11):2841-2850. DOI: 10.1111/ajt.14327.
27. Abdel-Hady Algharably E, Beige J, Kreutz R, Bolbrinker J. Effect of UMOD genotype on long-term graft survival after kidney transplantation in patients treated with cyclosporine-based therapy. Pharmacogenomics J 2018;18(2):227-231. DOI: 10.1038/tpj.2017.14.
28. Zhao D, Dai H, Macedo C, et al. Donor-recipient mismatch at the SIRPA locus adversely affects kidney allograft outcomes. Sci Transl Med 2025;17(807):eady1135. DOI: 10.1126/scitranslmed.ady1135.
29. Moore J, McKnight AJ, Dohler B, et al. Donor ABCB1 variant associates with increased risk for kidney allograft failure. J Am Soc Nephrol 2012;23(11):1891-9. DOI: 10.1681/ASN.2012030260.
30. Lanka S, K VR, Arji A, et al. Association of Tumor Necrosis Factor-Alpha (TNF-alpha) rs1800629 Polymorphism in Chronic Kidney Disease. Cureus 2024;16(5):e60332. DOI: 10.7759/cureus.60332.
31. Haas C, Ryffel B, Aguet M, Le Hir M. MHC antigens in interferon gamma (IFN gamma) receptor deficient mice: IFN gamma-dependent up-regulation of MHC class II in renal tubules. Kidney Int 1995;48(6):1721-7. DOI: 10.1038/ki.1995.470.
32. Saleh QW, Mohammadnejad A, Tepel M. FOXP3 full length splice variant is associated with kidney allograft tolerance. Front Immunol 2024;15:1389105. DOI: 10.3389/fimmu.2024.1389105.
33. Wanas H, Kamel MH, William EA, et al. The impact of CYP3A4 and CYP3A5 genetic variations on tacrolimus treatment of living-donor Egyptian kidney transplanted patients. J Clin Lab Anal 2023;37(19-20):e24969. DOI: 10.1002/jcla.24969.
34. Ozaltin F, Ibsirlioglu T, Taskiran EZ, et al. Disruption of PTPRO causes childhood-onset nephrotic syndrome. Am J Hum Genet 2011;89(1):139-47. DOI: 10.1016/j.ajhg.2011.05.026.
35. Ghisdal L, Baron C, Lebranchu Y, et al. Genome-Wide Association Study of Acute Renal Graft Rejection. Am J Transplant 2017;17(1):201-209. DOI: 10.1111/ajt.13912.
36. Isenberg JS, Roberts DD. The role of CD47 in pathogenesis and treatment of renal ischemia reperfusion injury. Pediatr Nephrol 2019;34(12):2479-2494. DOI: 10.1007/s00467-018-4123-z.
