## Supplementary Excel for "Quantifying donor-recipient mismatches using recipient-derived sources of donor DNA"

| Case-1 | | Rec AA and Donor BB | | | | Rec BB and Donor AA | | | | Rec AA and Donor AB | | | | Rec BB and Donor AB | | | | Rec AA and Donor | #Rec AA and Don | BB and Donor AA | Rec BB and Donor | AA and Donor AB | Rec AA and Donor | BB and Donor AB | BB and Donor AB | #Total MM | % Total MM | #Homo MM | % Homo MM | #Non-Detectable | %Non-Detectable |
|---|---|---|---|---|---|---|---|---|---|---|---|---|---|---|---|---|---|---|---|---|---|---|---|---|---|---|---|---|---|---|---|
| | | AA | AB | BB | None | AA | AB | BB | None | AA | AB | BB | None | AA | AB | BB | None | | | | | | | | | | | | | | |
| Whole Genome | | | | | | | | | | | | | | | | | | | | | | | | | | | | | | | |
| **Whole Genome** | **gDNA** | **6459** | | | | **7020** | | | | **44506** | | | | **13050** | | | | 6459 | | **7020** | | **44506** | | **13050** | | **71035** | | **13479** | | | |
| No HLA genes | cfDNA | 5363 | 33 | 6 | 1057 | 9 | 79 | 5785 | 1147 | 36348 | 990 | 3 | 7165 | 9 | 217 | 10701 | 2123 | **39** | 0.60 | **88** | 1.25 | **993** | 2.23 | **226** | 1.73 | 1346.00 | 1.89 | 15.00 | 0.11 | 11492 | 16.17794045 |
| | uDNA | 23 | 6097 | 277 | 62 | 462 | 6276 | 13 | 269 | 1332 | 42485 | 24 | 665 | 9 | 12799 | 176 | 66 | **6374** | 98.68 | **6738** | 95.98 | **42509** | 95.51 | **12808** | 98.15 | 68429.00 | 96.33 | 739.00 | 5.48 | 1062 | 1.495037657 |
| Secreted/Trans membrane | **gDNA** | **2935** | | | | **3250** | | | | **20119** | | | | **6032** | | | | **2935** | | **3250** | | 20119 | | 6032 | | **32336** | | **6185** | | | |
| | cfDNA | 2434 | 15 | 4 | 482 | 2 | 27 | 2724 | 497 | 16583 | 420 | 2 | 3114 | 8 | 95 | 5000 | 929 | **19** | 0.65 | **29** | 0.89 | **422** | 2.10 | **103** | 1.71 | 573 | 1.772018803 | 6.00 | 0.10 | 5022 | 15.53067788 |
| | uDNA | 12 | 2798 | 107 | 18 | 180 | 2962 | 8 | 100 | 447 | 19450 | 15 | 207 | 7 | 5928 | 72 | 25 | **2905** | 98.98 | **3142** | 96.68 | **19465** | 96.75 | **5935** | 98.39 | 31447 | 97.25074221 | 287.00 | 4.64 | 350 | 1.082384958 |
| **Exonic non-synonymous** | **gDNA** | **922** | | | | **951** | | | | **6074** | | | | **1782** | | | | **922** | | **951** | | **6074** | | **1782** | | **9729** | | **1873** | | | |
| Secreted/Trans membrane | cfDNA | 888 | 3 | 1 | 30 | 0 | 4 | 896 | 51 | 5724 | 91 | 0 | 259 | 1 | 22 | 1690 | 69 | **4** | 0.43 | **4** | 0.42 | **91** | 1.50 | **23** | 1.29 | 122 | 1.253982938 | 1.00 | 0.05 | 409 | 4.203926406 |
| | uDNA | 7 | 893 | 19 | 3 | 49 | 877 | 2 | 23 | 114 | 5921 | 1 | 38 | 0 | 1773 | 7 | 2 | **912** | 98.92 | **926** | 97.37 | **5922** | 97.50 | **1773** | 99.49 | 9533 | 97.98540446 | 68.00 | 3.63 | 66 | 0.678384212 |
| | **gDNA** | **152** | | | | **250** | | | | **1596** | | | | **572** | | | | **152** | | **250** | | **1596** | | **572** | | **2570** | | **402** | | | |
| Targeted Genes (56) | cfDNA | 137 | 2 | 1 | 12 | 1 | 2 | 225 | 22 | 1450 | 61 | 0 | 85 | 0 | 21 | 521 | 30 | **3** | 1.97 | **3** | 1.20 | **61** | 3.82 | **21** | 3.67 | 88 | 3.424124514 | 2.00 | 0.50 | 149 | 5.79766537 |
| | uDNA | 0 | 148 | 4 | 0 | 13 | 236 | 0 | 1 | 36 | 1540 | 3 | 17 | 0 | 551 | 21 | 0 | **152** | 100.00 | **249** | 99.60 | **1543** | 96.68 | **551** | 96.33 | 2495 | 97.08171206 | 17.00 | 4.23 | 18 | 0.700389105 |
| LIMS1 | **gDNA** | **0** | | | | **21** | | | | **171** | | | | **21** | | | | **0** | | **21** | | **171** | | **21** | | **213** | | **21** | | | |
| | cfDNA | 0 | 0 | 0 | 0 | 0 | 0 | 19 | 2 | 158 | 3 | 0 | 10 | 0 | 1 | 19 | 1 | **0** | 0.00 | **0** | 0.00 | **3** | | **1** | 4.76 | 4 | 1.877934272 | 0.00 | 0.00 | 13 | 6.103286385 |
| | uDNA | 0 | 0 | 0 | 0 | 0 | 21 | 0 | 0 | 1 | 170 | 0 | 0 | 0 | 19 | 2 | 0 | **0** | 0.00 | **21** | 100.00 | **170** | | **19** | 90.48 | 210 | 98.5915493 | 0.00 | 0.00 | 0 | 0 |

| Case-2 | | **Rec AA** and Donor BB | | | | Rec BB and Donor AA | | | | Rec AA and Donor AB | | | | Rec BB and Donor AB | | | | **A and Donor BB** | **mozygous misma** | **B and Donor AA** | **ec BB and Donor** | **A and Donor AB** | **ec AA and Donor** | **B and Donor AB** | **ec BB and Donor** | **Total** | **% Total** | **#Homo MM** | **% Homo MM** | #Non-Detectable | %Non-Detectable |
|---|---|---|---|---|---|---|---|---|---|---|---|---|---|---|---|---|---|---|---|---|---|---|---|---|---|---|---|---|---|---|---|
| | | AA | AB | BB | None | AA | AB | BB | None | AA | AB | BB | None | AA | AB | BB | None | | | | | | | | | | | | | | |
| Whole Genome | | | | | | | | | | | | | | | | | | | | | | | | | | | | | | | |
| Whole Genome | **gDNA** | **7389** | | | | **7120** | | | | **53535** | | | | **13808** | | | | **7389** | | 7120 | | | | | | **81852** | | **14509** | | | |
| No HLA genes | cfDNA | 5475 | 127 | 4 | 1783 | 12 | 333 | 5115 | 1660 | 39901 | 1057 | 5 | 12572 | 18 | 360 | 10149 | 3281 | **131** | 1.77 | 345 | 4.85 | 1062 | 1.98 | 378 | 2.74 | 1916.00 | 2.340810243 | 16.00 | 0.11 | 19296 | 23.57425597 |
| | uDNA | 483 | 6444 | 19 | 443 | 10 | 6561 | 356 | 193 | 22732 | 24779 | 13 | 6011 | 7 | 8523 | 4943 | 335 | **6463** | 87.47 | 6571 | 92.29 | 24792 | 46.31 | 8530 | 61.78 | 46356.00 | 56.63392464 | 29.00 | 0.20 | 6982 | 8.53002981 |
| | **gDNA** | **3413** | | | | **3144** | | | | **23790** | | | | **6316** | | | | **3413** | | 3144 | | | | | | **36663** | | **6557** | | | |
| Secreted/Trans membrane Genes | cfDNA | 2555 | 58 | 2 | 798 | 4 | 131 | 2284 | 725 | 17944 | 421 | 3 | 5422 | 8 | 160 | 4685 | 1463 | **60** | 1.76 | 135 | 4.29 | 424 | 1.78 | 168 | 2.66 | 787 | 2.146578294 | 6.00 | 0.09 | 8408 | 22.93320241 |
| | uDNA | 195 | 3025 | 9 | 184 | 6 | 2929 | 148 | 61 | 9880 | 11340 | 7 | 2563 | 4 | 3980 | 2213 | 119 | **3034** | 88.90 | 2935 | 93.35 | 11347 | 47.70 | 3984 | 63.08 | 21300 | 58.09671876 | 15.00 | 0.23 | 2927 | 7.983525625 |
| Exonic non-synonymous | **gDNA** | **981** | | | | **916** | | | | **7201** | | | | **1753** | | | | **981** | | 916 | | | | | | **10851** | | **1897** | | | |
| Secreted/Trans membrane | cfDNA | 897 | 12 | 1 | 71 | 2 | 22 | 837 | 55 | 6613 | 89 | 1 | 498 | 1 | 27 | 1594 | 131 | **13** | 1.33 | 24 | 2.62 | 90 | 1.98 | 28 | 1.60 | 155 | 1.428439775 | 3.00 | 0.16 | 755 | 6.957884066 |
| | uDNA | 44 | 908 | 0 | 29 | 0 | 863 | 35 | 18 | 3097 | 3495 | 2 | 607 | 1 | 1160 | 568 | 24 | **908** | 92.56 | 863 | 94.21 | 3497 | 46.31 | 1161 | 66.23 | 6429 | 59.24799558 | 0.00 | 0.00 | 678 | 6.248272049 |
| | **gDNA** | **142** | | | | **292** | | | | **2120** | | | | **670** | | | | **142** | | 292 | | | | | | **3224** | | **434** | | | |
| Targeted Genes (56) | cfDNA | 131 | 1 | 1 | 9 | 0 | 11 | 258 | 23 | 1923 | 57 | 1 | 139 | 1 | 31 | 596 | 42 | **2** | 1.41 | 11 | 3.77 | 58 | 2.74 | 32 | 4.78 | 103 | 3.194789082 | 1.00 | 0.23 | 213 | 6.606699752 |
| | uDNA | 5 | 132 | 2 | 3 | 0 | 289 | 2 | 1 | 860 | 1137 | 0 | 123 | 0 | 487 | 181 | 2 | **134** | 94.37 | 289 | 98.97 | 1137 | 53.63 | 487 | 72.69 | 2047 | 63.49255583 | 2.00 | 0.46 | 129 | 4.001240695 |
| | **gDNA** | **0** | | | | **1** | | | | **50** | | | | **11** | | | | **0** | | 0 | 0.00 | | | | | **62** | | **1** | | | |
| | cfDNA | 0 | 0 | 0 | 0 | 0 | 0 | 0 | 1 | 46 | 2 | 0 | 2 | 0 | 1 | 9 | 1 | **0** | 0.00 | 0 | 0.00 | 2 | 4.00 | 1 | 9.09 | 3 | 4.838709677 | 0.00 | 0.00 | 4 | 6.451612903 |
| LIMS1 | uDNA | 0 | 0 | 0 | 0 | 0 | 1 | 0 | 0 | 32 | 16 | 0 | 2 | 0 | 6 | 5 | 0 | **0** | 0.00 | 1 | 100.00 | 16 | 32.00 | 6 | 54.55 | 23 | 37.09677419 | 0.00 | 0.00 | 2 | 3.225806452 |

| Case-3 | | Rec AA and Donor BB | | | | Rec BB and Donor AA | | | | Rec AA and Donor AB | | | | Rec BB and Donor AB | | | | **A and Donor BB** | **mozygous misma** | **B and Donor AA** | **ec BB and Donor** | **A and Donor AB** | **ec AA and Donor** | **B and Donor AB** | **ec BB and Donor** | Total | % Total | #Homo MM | % Homo MM | #Non-Detectable | %Non-Detectable |
|---|---|---|---|---|---|---|---|---|---|---|---|---|---|---|---|---|---|---|---|---|---|---|---|---|---|---|---|---|---|---|---|
| | | AA | AB | BB | None | AA | AB | BB | None | AA | AB | BB | None | AA | AB | BB | None | | | | | | | | | | | | | | |
| Whole Genome | | | | | | | | | | | | | | | | | | | | | | | | | | | | | | | |
| Whole Genome | **gDNA** | **6649** | | | | **7792** | | | | **55552** | | | | **15291** | | | | **6649** | | **7792** | | **55552** | | **15291** | | **85284** | | **14441** | | | |
| No HLA genes | cfDNA | 4743 | 31 | 3 | 1872 | 21 | 64 | 5376 | 2331 | 39116 | 872 | 5 | 15559 | 14 | 193 | 10667 | 4417 | **34** | 0.51 | 85 | 1.09 | 877 | 1.58 | 207 | 1.35 | 1203 | 1.410581117 | 24.00 | 0.17 | 24179 | 28.35115614 |
| | uDNA | 0 | 0 | 0 | 0 | 0 | 0 | 0 | 0 | 0 | 0 | 0 | 0 | 0 | 0 | 0 | 0 | **0** | 0.00 | 0 | 0.00 | 0 | 0.00 | 0 | 0.00 | 0 | 0 | 0.00 | 0.00 | 0 | 0 |
| | **gDNA** | **3188** | | | | **3544** | | | | **24948** | | | | **6967** | | | | **3188** | | **3544** | | **24948** | | **6967** | | **38647** | | **6732** | | | |
| Secreted/Trans membrane Genes | cfDNA | 2302 | 15 | 0 | 871 | 10 | 21 | 2510 | 1003 | 17728 | 363 | 4 | 6853 | 6 | 92 | 4930 | 1939 | **15** | 0.47 | 31 | | 367 | 1.47 | 98 | 1.41 | 511 | 1.322224235 | 10.00 | 0.15 | 10666 | 27.59851994 |
| | uDNA | 0 | 0 | 0 | 0 | 0 | 0 | 0 | 0 | 0 | 0 | 0 | 0 | 0 | 0 | 0 | 0 | **0** | 0.00 | 0 | | 0 | 0.00 | 0 | 0.00 | 0 | 0 | 0.00 | 0.00 | 0 | 0 |
| Exonic non-synonymous | **gDNA** | **905** | | | | **1009** | | | | **7233** | | | | **1936** | | | | **905** | | 1009 | | 7233 | | 1936 | | **11083** | | **1914** | | | |
| Secreted/Trans membrane | cfDNA | 829 | 1 | 0 | 75 | 1 | 5 | 921 | 82 | 6529 | 104 | 0 | 600 | 0 | 19 | 1769 | 148 | **1** | 0.11 | 6 | 0.59 | 104 | 1.98 | 19 | 0.98 | 130 | 1.172967608 | 1.00 | 0.05 | 905 | 8.165659118 |
| | uDNA | 0 | 0 | 0 | 0 | 0 | 0 | 0 | 0 | 0 | 0 | 0 | 0 | 0 | 0 | 0 | 0 | **0** | 0.00 | 0 | 0.00 | 0 | 46.31 | 0 | 0.00 | 0 | 0 | 0.00 | 0.00 | 0 | 0 |
| | **gDNA** | **257** | | | | **268** | | | | **2810** | | | | **417** | | | | **257** | | 268 | | 2810 | | 417 | | **3752** | | **525** | | | |
| | cfDNA | 226 | 3 | 0 | 28 | 0 | 2 | 236 | 30 | 2502 | 48 | 2 | 258 | 1 | 11 | 359 | 46 | **3** | 1.17 | 2 | 0.75 | 50 | 1.78 | 12 | 2.88 | 67 | 1.785714286 | 0.00 | 0.00 | 362 | 9.648187633 |
| Targeted Genes (56) | uDNA | 0 | 0 | 0 | 0 | 0 | 0 | 0 | 0 | 0 | 0 | 0 | 0 | 0 | 0 | 0 | 0 | **0** | 0.00 | 0 | 0.00 | 0 | 0.00 | 0 | 0.00 | 0 | 0 | 0.00 | 0.00 | 0 | 0 |
| | **gDNA** | **0** | | | | **1** | | | | **86** | | | | **0** | | | | **0** | | 1 | | 86 | | 0 | | **87** | | **1** | | | |
| | cfDNA | 0 | 0 | 0 | 0 | 0 | 0 | 0 | 1 | 74 | 0 | 2 | 10 | 0 | 0 | 0 | 0 | **0** | 0.00 | 0 | 0.00 | 2 | 2.33 | 0 | #DIV/0! | 2 | 2.298850575 | 0.00 | 0.00 | 11 | 12.64367816 |
| LIMS1 | uDNA | 0 | 0 | 0 | 0 | 0 | 0 | 0 | 0 | 0 | 0 | 0 | 0 | 0 | 0 | 0 | 0 | **0** | 0.00 | 0 | 0.00 | 0 | 0.00 | 0 | #DIV/0! | 0 | 0 | 0.00 | 0.00 | 0 | 0 |

| Case-4 | | Rec AA and Donor BB | | | | Rec BB and Donor AA | | | | Rec AA and Donor AB | | | | Rec BB and Donor AB | | | | AA and Donor BB | ec AA and Donor | 3B and Donor AA | ec BB and Donor | AA and Donor AB | ec AA and Donor | 3B and Donor AB | ec BB and Donor | Total | % Total | #Homo MM | % Homo MM | #Non-Detectable | %Non-Detectable |
|---|---|---|---|---|---|---|---|---|---|---|---|---|---|---|---|---|---|---|---|---|---|---|---|---|---|---|---|---|---|---|---|
| | | AA | AB | BB | None | AA | AB | BB | None | AA | AB | BB | None | AA | AB | BB | None | | | | | | | | | | | | | | |
| Whole Genome | | | | | | | | | | | | | | | | | | | | | | | | | | | | | | | |
| Whole Genome | gDNA | 8112 | | | | 9325 | | | | 36434 | | | | 13636 | | | | 8112 | | 9325 | | 36434 | | 13636 | | 67507 | | 17437 | | | |
| No HLA genes | cfDNA | 6436 | 36 | 3 | 1637 | 18 | 75 | 7199 | 2033 | 27985 | 804 | 7 | 7638 | 9 | 206 | 10577 | 2844 | 39 | 0.48 | 93 | 1.00 | 811 | 2.23 | 215 | 1.58 | 1158 | 1.715377665 | 21.00 | 0.12 | 14152 | 20.96375191 |
| | uDNA | 17 | 6917 | 713 | 465 | 1146 | 7020 | 9 | 1150 | 1335 | 32733 | 21 | 2345 | 37 | 12703 | 162 | 734 | 7630 | 94.06 | 8166 | 87.57 | 32754 | 89.90 | 12740 | 93.43 | 61290 | 90.79058468 | 1859.00 | 10.66 | 4694 | 6.953352986 |
| | gDNA | 3631 | | | | 3976 | | | | 16480 | | | | 6404 | | | | 3631 | | 3976 | | 16480 | | 6404 | | 30491 | | 7607 | | | |
| Secreted/Trans membrane | cfDNA | 2885 | 14 | 2 | 730 | 5 | 31 | 3097 | 843 | 12938 | 324 | 5 | 3213 | 3 | 93 | 5067 | 1241 | 16 | 0.44 | 36 | 0.91 | 329 | 2.00 | 96 | 1.50 | 477 | 1.564396051 | 7.00 | 0.09 | 6027 | 19.76648847 |
| Genes | uDNA | 3 | 3125 | 319 | 184 | 453 | 3066 | 3 | 454 | 540 | 15145 | 7 | 788 | 13 | 6082 | 59 | 250 | 3444 | 94.85 | 3519 | 88.51 | 15152 | 91.94 | 6095 | 95.17 | 28210 | 92.519104 | 772.00 | 10.15 | 1676 | 5.496703945 |
| Exonic non-synonymous | gDNA | 1099 | | | | 1171 | | | | 4949 | | | | 1784 | | | | 1099 | | 1171 | | 4949 | | 1784 | | 9003 | | 2270 | | | |
| Secreted/Trans membrane | cfDNA | 1028 | 3 | 1 | 67 | 0 | 5 | 1096 | 70 | 4621 | 59 | 0 | 269 | 0 | 13 | 1641 | 130 | 4 | 0.36 | 5 | 0.43 | 59 | 1.98 | 13 | 0.73 | 81 | 0.8997001 | 1.00 | 0.04 | 536 | 5.953571032 |
| | uDNA | 0 | 986 | 76 | 37 | 115 | 971 | 1 | 84 | 129 | 4672 | 0 | 148 | 2 | 1726 | 10 | 46 | 1062 | 96.63 | 1086 | 92.74 | 4672 | 46.31 | 1728 | 96.86 | 8548 | 94.94612907 | 191.00 | 8.41 | 315 | 3.498833722 |
| | gDNA | 270 | | | | 243 | | | | 1995 | | | | 681 | | | | 270 | | 243 | | 1995 | | 681 | | 3189 | | 513 | | | |
| | cfDNA | 248 | 2 | 1 | 19 | 0 | 1 | 215 | 27 | 1806 | 48 | 1 | 140 | 1 | 14 | 619 | 47 | 3 | 1.11 | 1 | 0.41 | 49 | 2.46 | 15 | 2.20 | 68 | 2.132329884 | 1.00 | 0.19 | 233 | 7.306365632 |
| Targeted Genes (56) | uDNA | 0 | 254 | 12 | 4 | 19 | 212 | 0 | 12 | 41 | 1934 | 0 | 20 | 2 | 661 | 16 | 2 | 266 | 98.52 | 231 | 95.06 | 1934 | 96.94 | 663 | 97.36 | 3094 | 97.02100972 | 31.00 | 6.04 | 38 | 1.191596112 |
| | gDNA | 0 | | | | 3 | | | | 101 | | | | 51 | | | | 0 | | 3 | | 101 | | 51 | | 155 | | 3 | | | |
| | cfDNA | 0 | 0 | 0 | 0 | 0 | 0 | 0 | 3 | 92 | 2 | 0 | 7 | 1 | 0 | 46 | 4 | 0 | 0.00 | 0 | 0.00 | 2 | 1.98 | 1 | 1.96 | 3 | 1.935483871 | 0.00 | 0.00 | 14 | 9.032258065 |
| LIMS1 | uDNA | 0 | 0 | 0 | 0 | 1 | 1 | 0 | 1 | 5 | 93 | 0 | 3 | 0 | 50 | 1 | 0 | 0 | 0.00 | 2 | 66.67 | 93 | 92.08 | 50 | 98.04 | 145 | 93.5483871 | 1.00 | 33.33 | 4 | 2.580645161 |

| Case-5 | | Rec AA and Donor BB | | | | Rec BB and Donor AA | | | | Rec AA and Donor AB | | | | Rec BB and Donor AB | | | | AA and Donor BB | ec AA and Donor | c BB and Donor A | ec BB and Donor | c AA and Donor A | ec AA and Donor | c BB and Donor A | ec BB and Donor | Total | % Total | #Homo MM | % Homo MM | #Non-Detectable | %Non-Detectable |
|---|---|---|---|---|---|---|---|---|---|---|---|---|---|---|---|---|---|---|---|---|---|---|---|---|---|---|---|---|---|---|---|
| | | AA | AB | BB | None | AA | AB | BB | None | AA | AB | BB | None | AA | AB | BB | None | | | | | | | | | | | | | | |
| Whole Genome | | | | | | | | | | | | | | | | | | | | | | | | | | | | | | | |
| Whole Genome | gDNA | 12895 | | | | 9335 | | | | 63811 | | | | 13437 | | | | 12895 | | 9335 | | 63811 | | 13437 | | 99478 | | 22230 | | | |
| No HLA genes | cfDNA | 10916 | 114 | 8 | 1857 | 14 | 112 | 7811 | 1398 | 53139 | 1139 | 7 | 9526 | 7 | 233 | 11243 | 1954 | 122 | 0.95 | 126 | 1.35 | 1146 | 1.80 | 240 | 1.79 | 1634 | 1.642574238 | 22.00 | 0.10 | 14735 | 14.81232031 |
| | uDNA | 0 | 0 | 0 | 0 | 0 | 0 | 0 | 0 | 0 | 0 | 0 | 0 | 0 | 0 | 0 | 0 | 0 | 0.00 | 0 | 0.00 | 0 | 0.00 | 0 | 0.00 | 0 | 0 | 0.00 | 0.00 | 0 | 0 |
| | gDNA | 5958 | | | | 4363 | | | | 28523 | | | | 6136 | | | | 5958 | | 4363 | | 28523 | | 6136 | | 44980 | | 10321 | | | |
| Secreted/Trans membrane | cfDNA | 5093 | 56 | 4 | 805 | 6 | 60 | 3668 | 629 | 23910 | 465 | 5 | 4143 | 3 | 82 | 5200 | 851 | 60 | 1.01 | 66 | 1.51 | 470 | 1.65 | 85 | 1.39 | 681 | 1.514006225 | 10.00 | 0.10 | 6428 | 14.29079591 |
| Genes | uDNA | 0 | 0 | 0 | 0 | 0 | 0 | 0 | 0 | 0 | 0 | 0 | 0 | 0 | 0 | 0 | 0 | 0 | 0.00 | 0 | 0.00 | 0 | 0.00 | 0 | 0.00 | 0 | 0 | 0.00 | 0.00 | 0 | 0 |
| Exonic non-synonymous | gDNA | 1802 | | | | 1268 | | | | 8134 | | | | 1664 | | | | 1802 | | 1268 | | 8134 | | 1664 | | 12868 | | 3070 | | | |
| Secreted/Trans membrane | cfDNA | 1695 | 13 | 0 | 94 | 0 | 15 | 1196 | 57 | 7696 | 74 | 0 | 364 | 0 | 5 | 1585 | 74 | 13 | 0.72 | 15 | 1.18 | 74 | 1.98 | 5 | 0.30 | 107 | 0.83152005 | 0.00 | 0.00 | 589 | 4.577245881 |
| | uDNA | 0 | 0 | 0 | 0 | 0 | 0 | 0 | 0 | 0 | 0 | 0 | 0 | 0 | 0 | 0 | 0 | 0 | 0.00 | 0 | 0.00 | 0 | 46.31 | 0 | 0.00 | 0 | 0 | 0.00 | 0.00 | 0 | 0 |
| | gDNA | 509 | | | | 316 | | | | 2760 | | | | 606 | | | | 509 | | 316 | | 2760 | | 606 | | 4191 | | 825 | | | |
| | cfDNA | 489 | 7 | 0 | 13 | 0 | 10 | 294 | 12 | 2609 | 72 | 0 | 79 | 0 | 16 | 572 | 18 | 7 | 1.38 | 10 | 3.16 | 72 | 2.61 | 16 | 2.64 | 105 | 2.505368647 | 0.00 | 0.00 | 122 | 2.910999761 |
| Targeted Genes (56) | uDNA | 0 | 0 | 0 | 0 | 0 | 0 | 0 | 0 | 0 | 0 | 0 | 0 | 0 | 0 | 0 | 0 | 0 | 0.00 | 0 | 0.00 | 0 | 0.00 | 0 | 0.00 | 0 | 0 | 0.00 | 0.00 | 0 | 0 |
| | gDNA | 1 | | | | 0 | | | | 50 | | | | 33 | | | | 1 | | 0 | | 50 | | 33 | | 84 | | 1 | | | |
| | cfDNA | 0 | 1 | 0 | 0 | 0 | 0 | 0 | 0 | 41 | 4 | 0 | 5 | 0 | 1 | 32 | 0 | 1 | 0.00 | 0 | #DIV/0! | 4 | 8.00 | 1 | 3.03 | 6 | 7.142857143 | 0.00 | 0.00 | 5 | 5.952380952 |
| LIMS1 | uDNA | 0 | 0 | 0 | 0 | 0 | 0 | 0 | 0 | 0 | 0 | 0 | 0 | 0 | 0 | 0 | 0 | 0 | 0.00 | 0 | #DIV/0! | 0 | 0.00 | 0 | 0.00 | 0 | 0 | 0.00 | 0.00 | 0 | 0 |

| Case-6 | | Rec AA and Donor BB | | | | Rec BB and Donor AA | | | | Rec AA and Donor AB | | | | Rec BB and Donor AB | | | | AA and Donor BB | ec AA and Donor | 3B and Donor AA | ec BB and Donor | AA and Donor AB | ec AA and Donor | 3B and Donor AB | ec BB and Donor | Total | % Total | #Homo MM | % Homo MM | #Non-Detectable | %Non-Detectable |
|---|---|---|---|---|---|---|---|---|---|---|---|---|---|---|---|---|---|---|---|---|---|---|---|---|---|---|---|---|---|---|---|
| | | AA | AB | BB | None | AA | AB | BB | None | AA | AB | BB | None | AA | AB | BB | None | | | | | | | | | | | | | | |
| | cfDNA | | | | | | | | | | | | | | | | | | | | | | | | | | | | | | |
| Whole Genome | uDNA | | | | | | | | | | | | | | | | | | | | | | | | | | | | | | |
| Whole Genome | | 6180 | | | | 9839 | | | | 48569 | | | | 16333 | | | | 6180 | | 9839 | | 48569 | | 16333 | | 80921 | | 16019 | | | |
| No HLA genes | cfDNA | 5011 | 45 | 5 | 1119 | 24 | 94 | 8012 | 1709 | 39368 | 808 | 4 | 8389 | 14 | 229 | 13343 | 2747 | 50 | 0.81 | 118 | 1.20 | 812 | 1.67 | 243 | 1.49 | 1223 | 1.511350576 | 29.00 | 0.18 | 13964 | 17.25633643 |
| | uDNA | 0 | 0 | 0 | 0 | 0 | 0 | 0 | 0 | 0 | 0 | 0 | 0 | 0 | 0 | 0 | 0 | 0 | 0.00 | 0 | 0.00 | 0 | 0.00 | 0 | 0.00 | 0 | 0 | 0.00 | 0.00 | 0 | 0 |
| | | 2702 | | | | 4300 | | | | 22114 | | | | 7426 | | | | 2702 | | 4300 | | 22114 | | 7426 | | 36542 | | 7002 | | | |
| Secreted/Trans membrane | cfDNA | 2208 | 18 | 3 | 473 | 17 | 36 | 3530 | 717 | 18066 | 345 | 1 | 3702 | 6 | 102 | 6089 | 1229 | 21 | 0.78 | 53 | 1.23 | 346 | 1.56 | 108 | 1.45 | 528 | 1.444912703 | 20.00 | 0.29 | 6121 | 16.75058836 |
| Genes | uDNA | 0 | 0 | 0 | 0 | 0 | 0 | 0 | 0 | 0 | 0 | 0 | 0 | 0 | 0 | 0 | 0 | 0 | 0.00 | 0 | 0.00 | 0 | 0.00 | 0 | 0.00 | 0 | 0 | 0.00 | 0.00 | 0 | 0 |
| Exonic non-synonymous | | 794 | | | | 1336 | | | | 6844 | | | | 2095 | | | | 794 | | 1336 | | 6844 | | 2095 | | 11069 | | 2130 | | | |
| Secreted/Trans membrane | cfDNA | 764 | 2 | 1 | 27 | 2 | 8 | 1256 | 70 | 6443 | 68 | 0 | 333 | 2 | 19 | 1969 | 105 | 3 | 0.38 | 10 | 0.75 | 68 | 1.98 | 21 | 1.00 | 102 | 0.921492456 | 3.00 | 0.14 | 535 | 4.833318276 |
| | uDNA | 0 | 0 | 0 | 0 | 0 | 0 | 0 | 0 | 0 | 0 | 0 | 0 | 0 | 0 | 0 | 0 | 0 | 0.00 | 0 | 0.00 | 0 | 46.31 | 0 | 0.00 | 0 | 0 | 0.00 | 0.00 | 0 | 0 |
| | | 217 | | | | 546 | | | | 1972 | | | | 1004 | | | | 217 | | 546 | | 1972 | | 1004 | | 3739 | | 763 | | | |
| | cfDNA | 206 | 3 | 0 | 8 | 0 | 7 | 519 | 20 | 1808 | 51 | 1 | 112 | 0 | 27 | 915 | 62 | 3 | 1.38 | 7 | 1.28 | 52 | 2.64 | 27 | 2.69 | 89 | 2.380315592 | 0.00 | 0.00 | 202 | 5.402514041 |
| Targeted Genes (56) | uDNA | 0 | 0 | 0 | 0 | 0 | 0 | 0 | 0 | 0 | 0 | 0 | 0 | 0 | 0 | 0 | 0 | 0 | 0.00 | 0 | 0.00 | 0 | 0.00 | 0 | 0.00 | 0 | 0 | 0.00 | 0.00 | 0 | 0 |
| | | 0 | | | | 1 | | | | 61 | | | | 45 | | | | 0 | | 1 | | 61 | | 45 | | 107 | | 1 | | | |
| | cfDNA | 0 | 0 | 0 | 0 | 0 | 0 | 1 | 0 | 49 | 4 | 0 | 8 | 0 | 1 | 44 | 0 | 0 | 0.00 | 0 | 0.00 | 4 | 6.56 | 1 | 2.22 | 5 | 4.672897196 | 0.00 | 0.00 | 8 | 7.476635514 |
| LIMS1 | uDNA | 0 | 0 | 0 | 0 | 0 | 0 | 0 | 0 | 0 | 0 | 0 | 0 | 0 | 0 | 0 | 0 | 0 | 0.00 | 0 | 0.00 | 0 | 0.00 | 0 | 0.00 | 0 | 0 | 0.00 | 0.00 | 0 | 0 |

| Case-8 | | Rec AA and Donor BB | | | | Rec BB and Donor AA | | | | Rec AA and Donor AB | | | | Rec BB and Donor AB | | | | AA and Donor BB | ec AA and Donor | 3B and Donor AA | ec BB and Donor | A and Donor AB | ec AA and Donor | 3B and Donor AB | ec BB and Donor | Total | % Total | #Homo MM | % Homo MM | #Non-Detectable | %Non-Detectable |
|---|---|---|---|---|---|---|---|---|---|---|---|---|---|---|---|---|---|---|---|---|---|---|---|---|---|---|---|---|---|---|---|
| | | AA | AB | BB | None | AA | AB | BB | None | AA | AB | BB | None | AA | AB | BB | None | | | | | | | | | | | | | | |
| Whole Genome | | | | | | | | | | | | | | | | | | | | | | | | | | | | | | | |
| Whole Genome | gDNA | 6197 | | | | 6490 | | | | 34197 | | | | 11862 | | | | 6197 | | 6490 | | 34197 | | 11862 | | 58746 | | 12687 | | | |
| No HLA genes | cfDNA | 5049 | 40 | 9 | 1099 | 16 | 45 | 5224 | 1205 | 27396 | 984 | 7 | 5810 | 10 | 170 | 9604 | 2078 | 49 | 0.79 | 61 | 0.94 | 991 | 2.90 | 180 | 1.52 | 1281 | 2.180573997 | 25.00 | 0.20 | 10192 | 17.34926633 |
| | uDNA | 8 | 5461 | 667 | 61 | 928 | 5031 | 16 | 515 | 1293 | 32388 | 32 | 484 | 10 | 11644 | 134 | 74 | 6128 | 98.89 | 5959 | 91.82 | 32420 | 94.80 | 11654 | 98.25 | 56161 | 95.59970041 | 1595.00 | 12.57 | 1134 | 1.930344194 |
| | gDNA | 2828 | | | | 2988 | | | | 15279 | | | | 5438 | | | | 2828 | | 2988 | | 15279 | | 5438 | | 26533 | | 5816 | | | |
| Secreted/Trans membrane | cfDNA | 2298 | 19 | 2 | 509 | 10 | 22 | 2418 | 538 | 12313 | 388 | 2 | 2576 | 5 | 78 | 4418 | 937 | 21 | 0.74 | 32 | 1.07 | 390 | 2.55 | 83 | 1.53 | 526 | 1.982436965 | 12.00 | 0.21 | 4560 | 17.18614555 |
| Genes | uDNA | 4 | 2520 | 286 | 18 | 391 | 2363 | 10 | 224 | 483 | 14618 | 6 | 172 | 4 | 5348 | 61 | 25 | 2806 | 99.22 | 2754 | 92.17 | 14624 | 95.71 | 5352 | 98.42 | 25536 | 96.24241511 | 677.00 | 11.64 | 439 | 1.654543399 |
| Exonic non-synonymous | gDNA | 859 | | | | 946 | | | | 4701 | | | | 1649 | | | | 859 | | 946 | | 4701 | | 1649 | | 8155 | | 1805 | | | |
| Secreted/Trans membrane | cfDNA | 815 | 0 | 1 | 43 | 1 | 5 | 905 | 35 | 4379 | 108 | 0 | 214 | 0 | 9 | 1557 | 83 | 1 | 0.12 | 6 | 0.63 | 108 | 1.98 | 9 | 0.55 | 124 | 1.520539546 | 2.00 | 0.11 | 375 | 4.598405886 |
| | uDNA | 1 | 815 | 39 | 4 | 113 | 790 | 1 | 42 | 120 | 4541 | 0 | 40 | 1 | 1640 | 5 | 3 | 854 | 99.42 | 903 | 95.45 | 4541 | 46.31 | 1641 | 99.51 | 7939 | 97.35131821 | 152.00 | 8.42 | 89 | 1.091354997 |
| | gDNA | 147 | | | | 164 | | | | 1502 | | | | 383 | | | | 147 | | 164 | | 1502 | | 383 | | 2196 | | 311 | | | |
| Targeted Genes (56) | cfDNA | 127 | 6 | 1 | 13 | 0 | 3 | 145 | 16 | 1365 | 67 | 1 | 69 | 0 | 16 | 340 | 27 | 7 | 4.76 | 3 | 1.83 | 68 | 4.53 | 16 | 4.18 | 94 | 4.280510018 | 1.00 | 0.32 | 125 | 5.692167577 |
| | uDNA | 0 | 132 | 13 | 2 | 16 | 141 | 1 | 6 | 29 | 1469 | 1 | 3 | 1 | 376 | 5 | 1 | 145 | 98.64 | 157 | 95.73 | 1470 | 97.87 | 377 | 98.43 | 2149 | 97.85974499 | 29.00 | 9.32 | 12 | 0.546448087 |
| | gDNA | 1 | | | | 0 | | | | 21 | | | | 1 | | | | 1 | | 0 | | 21 | | 1 | | 23 | | 1 | | | |
| | cfDNA | 1 | 0 | 0 | 0 | 0 | 0 | 0 | 0 | 18 | 2 | 0 | 1 | 0 | 0 | 0 | 1 | 0 | 0.00 | 0 | #DIV/0! | 2 | 9.52 | 0 | 0.00 | 2 | 8.695652174 | 0.00 | 0.00 | 2 | 8.695652174 |
| LIMS1 | uDNA | 0 | 0 | 1 | 0 | 0 | 0 | 0 | 0 | 2 | 19 | 0 | 0 | 1 | 0 | 0 | 0 | 1 | 0.00 | 0 | #DIV/0! | 19 | 90.48 | 1 | 100.00 | 21 | 91.30434783 | 1.00 | 100.00 | 0 | 0 |

| Case-9 | | Rec AA and Donor BB | | | | Rec BB and Donor AA | | | | Rec AA and Donor AB | | | | Rec BB and Donor AB | | | | AA and Donor BB | ec AA and Donor | 3B and Donor AA | ec BB and Donor | A and Donor AB | ec AA and Donor | 3B and Donor AB | ec BB and Donor | Total | % Total | #Homo MM | % Homo MM | #Non-Detectable | %Non-Detectable |
|---|---|---|---|---|---|---|---|---|---|---|---|---|---|---|---|---|---|---|---|---|---|---|---|---|---|---|---|---|---|---|---|
| | | AA | AB | BB | None | AA | AB | BB | None | AA | AB | BB | None | AA | AB | BB | None | | | | | | | | | | | | | | |
| Whole Genome | | | | | | | | | | | | | | | | | | | | | | | | | | | | | | | |
| Whole Genome | gDNA | 6031 | | | | 6214 | | | | 34136 | | | | 11905 | | | | 6031 | | 6214 | | 34136 | | 11905 | | 58286 | | 12245 | | | |
| No HLA genes | cfDNA | 5024 | 43 | 3 | 961 | 13 | 50 | 5114 | 1037 | 27328 | 1077 | 12 | 5719 | 10 | 209 | 9776 | 1910 | 46 | 0.76 | 63 | 1.01 | 1089 | 3.19 | 219 | 1.84 | 1417 | 2.431115534 | 16.00 | 0.13 | 9627 | 16.5168308 |
| | uDNA | 21 | 5802 | 35 | 173 | 50 | 5965 | 11 | 188 | 1637 | 31108 | 14 | 1377 | 6 | 11344 | 260 | 295 | 5837 | 96.78 | 6015 | 96.80 | 31122 | 91.17 | 11350 | 95.34 | 54324 | 93.2024843 | 85.00 | 0.69 | 2033 | 3.487973098 |
| | gDNA | 2799 | | | | 2823 | | | | 15285 | | | | 5400 | | | | 2799 | | 2823 | | 15285 | | 5400 | | 26307 | | 5622 | | | |
| Secreted/Trans membrane | cfDNA | 2358 | 20 | 0 | 421 | 5 | 23 | 2335 | 460 | 12372 | 462 | 8 | 2443 | 3 | 73 | 4460 | 864 | 20 | 0.71 | 28 | 0.99 | 470 | 3.07 | 76 | 1.41 | 594 | 2.257954157 | 5.00 | 0.09 | 4188 | 15.91971719 |
| Genes | uDNA | 6 | 2721 | 13 | 59 | 15 | 2728 | 6 | 74 | 627 | 14166 | 5 | 487 | 3 | 5188 | 94 | 115 | 2734 | 97.68 | 2743 | 97.17 | 14171 | 92.71 | 5191 | 96.13 | 24839 | 94.41973619 | 28.00 | 0.50 | 735 | 2.793933174 |
| Exonic non-synonymous | gDNA | 835 | | | | 828 | | | | 4727 | | | | 1557 | | | | 835 | | 828 | | 4727 | | 1557 | | 7947 | | 1663 | | | |
| Secreted/Trans membrane | cfDNA | 792 | 5 | 0 | 38 | 0 | 6 | 772 | 50 | 4391 | 105 | 1 | 230 | 0 | 15 | 1470 | 72 | 5 | 0.60 | 6 | 0.72 | 106 | 1.98 | 15 | 0.96 | 132 | 1.661004153 | 0.00 | 0.00 | 390 | 4.907512269 |
| | uDNA | 0 | 823 | 4 | 8 | 6 | 810 | 0 | 12 | 160 | 4489 | 2 | 76 | 0 | 1538 | 10 | 9 | 827 | 99.04 | 816 | 98.55 | 4491 | 46.31 | 1538 | 98.78 | 7672 | 96.53957468 | 10.00 | 0.60 | 105 | 1.321253303 |
| | gDNA | 429 | | | | 479 | | | | 1216 | | | | 563 | | | | 429 | | 479 | | 1216 | | 563 | | 2687 | | 908 | | | |
| | cfDNA | 406 | 3 | 0 | 20 | 1 | 6 | 452 | 20 | 1090 | 57 | 3 | 66 | 0 | 22 | 520 | 21 | 3 | 0.70 | 7 | 1.46 | 60 | 4.93 | 22 | 3.91 | 92 | 3.423892817 | 1.00 | 0.11 | 127 | 4.726460737 |
| Targeted Genes (56) | uDNA | 2 | 423 | 1 | 3 | 2 | 477 | 0 | 0 | 44 | 1156 | 1 | 15 | 0 | 548 | 15 | 0 | 424 | 98.83 | 479 | 100.00 | 1157 | 95.15 | 548 | 97.34 | 2608 | 97.05991812 | 3.00 | 0.33 | 18 | 0.669892073 |
| | gDNA | 2 | | | | 0 | | | | 14 | | | | 2 | | | | 2 | | 0 | | 14 | | 2 | | 18 | | 2 | | | |
| | cfDNA | 1 | 0 | 0 | 1 | 0 | 0 | 0 | 0 | 12 | 1 | 0 | 1 | 0 | 1 | 0 | 1 | 0 | 0.00 | 0 | #DIV/0! | 1 | 7.14 | 1 | 50.00 | 2 | 11.11111111 | 0.00 | 0.00 | 3 | 16.66666667 |
| LIMS1 | uDNA | 0 | 1 | 1 | 0 | 0 | 0 | 0 | 0 | 4 | 10 | 0 | 0 | 0 | 1 | 1 | 0 | 2 | 0.00 | 0 | #DIV/0! | 10 | 71.43 | 1 | 50.00 | 13 | 72.22222222 | 1.00 | 50.00 | 0 | 0 |

| Case-10 | | Rec AA and Donor BB | | | | Rec BB and Donor AA | | | | Rec AA and Donor AB | | | | Rec BB and Donor AB | | | | AA and Donor BB | ec AA and Donor | 3B and Donor AA | ec BB and Donor | A and Donor AB | ec AA and Donor | 3B and Donor AB | ec BB and Donor | Total | % Total | #Homo MM | % Homo MM | #Non-Detectable | %Non-Detectable |
|---|---|---|---|---|---|---|---|---|---|---|---|---|---|---|---|---|---|---|---|---|---|---|---|---|---|---|---|---|---|---|---|
| | | AA | AB | BB | None | AA | AB | BB | None | AA | AB | BB | None | AA | AB | BB | None | | | | | | | | | | | | | | |
| Whole Genome | | | | | | | | | | | | | | | | | | | | | | | | | | | | | | | |
| Whole Genome | gDNA | 5908 | | | | 5501 | | | | 35719 | | | | 12324 | | | | 5908 | | 5501 | | 35719 | | 12324 | | 59452 | | 11409 | | | |
| No HLA genes | cfDNA | 4552 | 56 | 2 | 1298 | 5 | 135 | 4090 | 1271 | 26794 | 939 | 8 | 7978 | 14 | 241 | 9321 | 2748 | 58 | 0.98 | 140 | 2.54 | 947 | 2.65 | 255 | 2.07 | 1400 | 2.35484088 | 7.00 | 0.06 | 13295 | 22.36257821 |
| | uDNA | 5027 | 70 | 3 | 808 | 10 | 98 | 4560 | 833 | 29520 | 1298 | 15 | 4886 | 9 | 279 | 10308 | 1728 | 73 | 1.24 | 108 | 1.96 | 1313 | 3.68 | 288 | 2.34 | 1782 | 2.997376034 | 13.00 | 0.11 | 8255 | 13.88515105 |
| | gDNA | 2659 | | | | 2433 | | | | 16319 | | | | 5620 | | | | 2659 | | 2433 | | 16319 | | 5620 | | 27031 | | 5092 | | | |
| Secreted/Trans membrane | cfDNA | 2069 | 25 | 0 | 565 | 1 | 69 | 1827 | 536 | 12381 | 391 | 3 | 3544 | 5 | 103 | 4279 | 1233 | 25 | 0.94 | 70 | 2.88 | 394 | 2.41 | 108 | 1.92 | 597 | 2.208575339 | 1.00 | 0.02 | 5878 | 21.74540343 |
| Genes | uDNA | 2277 | 35 | 2 | 345 | 3 | 48 | 2023 | 359 | 13532 | 571 | 7 | 2209 | 5 | 128 | 4709 | 778 | 37 | 1.39 | 51 | 2.10 | 578 | 3.54 | 133 | 2.37 | 799 | 2.955865488 | 5.00 | 0.10 | 3691 | 13.65469276 |
| Exonic non-synonymous | gDNA | 770 | | | | 709 | | | | 4978 | | | | 1613 | | | | 770 | | 709 | | 4978 | | 1613 | | 8070 | | 1479 | | | |
| Secreted/Trans membrane | cfDNA | 732 | 4 | 0 | 34 | 1 | 19 | 642 | 47 | 4550 | 84 | 1 | 343 | 1 | 14 | 1508 | 90 | 4 | 0.52 | 20 | 2.82 | 85 | 1.98 | 15 | 0.93 | 124 | 1.536555143 | 1.00 | 0.07 | 514 | 6.369268897 |
| | uDNA | 731 | 6 | 0 | 33 | 2 | 11 | 656 | 40 | 4594 | 99 | 0 | 285 | 0 | 16 | 1518 | 79 | 6 | 0.78 | 13 | 1.83 | 99 | 46.31 | 16 | 0.99 | 134 | 1.66047088 | 2.00 | 0.14 | 437 | 5.41511772 |
| | gDNA | 295 | | | | 410 | | | | 1486 | | | | 611 | | | | 295 | | 410 | | 1486 | | 611 | | 2802 | | 705 | | | |
| | cfDNA | 282 | 6 | 0 | 7 | 0 | 29 | 350 | 31 | 1346 | 49 | 0 | 91 | 2 | 9 | 553 | 47 | 6 | 2.03 | 29 | 7.07 | 49 | 3.30 | 11 | 1.80 | 95 | 3.390435403 | 0.00 | 0.00 | 176 | 6.281227695 |
| Targeted Genes (56) | uDNA | 283 | 5 | 0 | 7 | 0 | 7 | 381 | 22 | 1358 | 79 | 0 | 49 | 0 | 15 | 581 | 15 | 5 | 1.69 | 7 | 1.71 | 79 | 5.32 | 15 | 2.45 | 106 | 3.783012134 | 0.00 | 0.00 | 93 | 3.319057816 |
| | gDNA | 1 | | | | 0 | | | | 11 | | | | 1 | | | | 1 | | 0 | | 11 | | 1 | | 13 | | 1 | | | |
| | cfDNA | 1 | 0 | 0 | 0 | 0 | 0 | 0 | 0 | 7 | 2 | 0 | 2 | 0 | 0 | 1 | 0 | 0 | 0.00 | 0 | #DIV/0! | 2 | 18.18 | 0 | 0.00 | 2 | 15.38461538 | 0.00 | 0.00 | 2 | 15.38461538 |
| LIMS1 | uDNA | 1 | 0 | 0 | 0 | 0 | 0 | 0 | 0 | 6 | 5 | 0 | 0 | 0 | 1 | 0 | 0 | 0 | 0.00 | 0 | #DIV/0! | 5 | 45.45 | 1 | 100.00 | 6 | 46.15384615 | 0.00 | 0.00 | 0 | 0 |

| Case-11 | | Rec AA and Donor BB | | | | Rec BB and Donor AA | | | | Rec AA and Donor AB | | | | Rec BB and Donor AB | | | | A and Donor BB | ec AA and Donor | B and Donor AA | ec BB and Donor | A and Donor AB | ec AA and Donor | B and Donor AB | ec BB and Donor | #Total | % Total | #Homo MM | % Homo MM | #Non-Detectable | %Non-Detectable |
|---|---|---|---|---|---|---|---|---|---|---|---|---|---|---|---|---|---|---|---|---|---|---|---|---|---|---|---|---|---|---|---|
| | | AA | AB | BB | None | AA | AB | BB | None | AA | AB | BB | None | AA | AB | BB | None | | | | | | | | | | | | | | |
| Whole Genome | | | | | | | | | | | | | | | | | | | | | | | | | | | | | | | |
| Whole Genome | **gDNA** | **9436** | | | | **11885** | | | | **39662** | | | | **13365** | | | | **9436** | | 11885 | | 39662 | | 13365 | | **74348** | | **21321** | | | |
| No HLA genes | cfDNA | 7524 | 48 | 1 | 1863 | 14 | 91 | 9314 | 2466 | 30733 | 795 | 6 | 8128 | 13 | 204 | 10446 | 2702 | **49** | 0.52 | 105 | 0.88 | 801 | 2.02 | 217 | 1.62 | 1172 | 1.576370582 | 15.00 | 0.07 | 15159 | 20.38925055 |
| | uDNA | 26 | 9213 | 76 | 121 | 159 | 11489 | 13 | 224 | 1553 | 37269 | 21 | 819 | 6 | 12905 | 282 | 172 | **9289** | 98.44 | 11648 | 98.01 | 37290 | 94.02 | 12911 | 96.60 | 71138 | 95.68246624 | 235.00 | 1.10 | 1336 | 1.796954861 |
| | **gDNA** | **4037** | | | | **5448** | | | | **17708** | | | | **5997** | | | | **4037** | | 5448 | | 17708 | | 5997 | | **33190** | | **9485** | | | |
| Secreted/Trans membrane | cfDNA | 3208 | 14 | 0 | 815 | 6 | 40 | 4315 | 1087 | 13872 | 339 | 3 | 3494 | 9 | 94 | 4729 | 1165 | **14** | 0.35 | 46 | 0.84 | 342 | 1.93 | 103 | 1.72 | 505 | 1.521542633 | 6.00 | 0.06 | 6561 | 19.76800241 |
| | uDNA | 6 | 3963 | 33 | 35 | 40 | 5313 | 5 | 90 | 534 | 16907 | 8 | 259 | 2 | 5812 | 126 | 57 | **3996** | 98.98 | 5353 | 98.26 | 16915 | 95.52 | 5814 | 96.95 | 32078 | 96.64959325 | 73.00 | 0.77 | 441 | 1.328713468 |
| | **gDNA** | **1195** | | | | **1566** | | | | **5371** | | | | **1677** | | | | **1195** | | 1566 | | 5371 | | 1677 | | **9809** | | **2761** | | | |
| Exonic non-synonymous | cfDNA | 1121 | 5 | 0 | 69 | 1 | 4 | 1459 | 102 | 5030 | 69 | 1 | 271 | 0 | 14 | 1562 | 101 | **5** | 0.42 | 5 | 0.32 | 70 | 1.98 | 14 | 0.83 | 94 | 0.958303599 | 1.00 | 0.04 | 543 | 5.535732491 |
| Secreted/Trans membrane | uDNA | 0 | 1184 | 7 | 4 | 8 | 1543 | 1 | 14 | 139 | 5193 | 0 | 39 | 0 | 1659 | 14 | 4 | **1191** | 99.67 | 1551 | 99.04 | 5193 | 46.31 | 1659 | 98.93 | 9594 | 97.80813539 | 15.00 | 0.54 | 61 | 0.621877867 |
| | | **201** | | | | **289** | | | | **1745** | | | | **637** | | | | **201** | | 289 | | 1745 | | 637 | | **2872** | | **490** | | | |
| | cfDNA | 187 | 3 | 0 | 11 | 0 | 1 | 264 | 24 | 1595 | 45 | 2 | 103 | 1 | 19 | 580 | 37 | **3** | 1.49 | 1 | 0.35 | 47 | 2.69 | 20 | 3.14 | 71 | 2.472144847 | 0.00 | 0.00 | 175 | 6.093314763 |
| Targeted Genes (56) | uDNA | 2 | 194 | 0 | 5 | 0 | 286 | 1 | 2 | 43 | 1689 | 0 | 13 | 0 | 614 | 23 | 0 | **194** | 96.52 | 286 | 98.96 | 1689 | 96.79 | 614 | 96.39 | 2783 | 96.90111421 | 0.00 | 0.00 | 20 | 0.69637883 |
| | **gDNA** | **2** | | | | **1** | | | | **130** | | | | **14** | | | | **2** | | 1 | | 130 | | 14 | | **147** | | **3** | | | |
| | cfDNA | 2 | 0 | 0 | 0 | 0 | 0 | 0 | 1 | 122 | 4 | 0 | 4 | 0 | 1 | 12 | 1 | **0** | 0.00 | 0 | 0.00 | 4 | 3.08 | 1 | 7.14 | 5 | 3.401360544 | 0.00 | 0.00 | 6 | 4.081632653 |
| LIMS1 | uDNA | 0 | 2 | 0 | 0 | 0 | 1 | 0 | 0 | 3 | 126 | 0 | 1 | 0 | 13 | 1 | 0 | **2** | 0.00 | 1 | 100.00 | 126 | 96.92 | 13 | 92.86 | 142 | 96.59863946 | 0.00 | 0.00 | 1 | 0.680272109 |

| Control | | Rec AA | | | | Rec BB | | | | Rec AB | | | | Total | (%) | al Non-datecton-Detectable | |
|---|---|---|---|---|---|---|---|---|---|---|---|---|---|---|---|---|---|
| | | AA | AB | BB | None | AA | AB | BB | None | AA | AB | BB | None | | | | |
| Whole Exome | gDNA | **92007** | | | | **58631** | | | | **103886** | | | | 254524 | **254524** | **254524** | |
| No HLA genes | cfDNA | 61003 | 13494 | 4 | 17506 | 1 | 346 | 54586 | 3698 | 4765 | 92680 | 312 | 6129 | **18922** | **7.4342695** | 27333 | 10.73887 |
| | uDNA | 71526 | 4824 | 2 | 15655 | 4 | 1555 | 55231 | 1841 | 4011 | 94878 | 237 | 4760 | **10633** | **4.1776021** | 22256 | 8.744166 |
| | gDNA | **7142** | | | | **10314** | | | | **15448** | | | | 32904 | **32904** | **32904** | |
| /Transmembran | cfDNA | 6007 | 107 | 0 | 1028 | 0 | 52 | 9688 | 574 | 177 | 14585 | 50 | 636 | **386** | **1.1731097** | 2238 | 6.801605 |
| | uDNA | 5460 | 210 | 0 | 1472 | 0 | 239 | 9791 | 284 | 125 | 14775 | 34 | 514 | **608** | **1.8477997** | 2270 | 6.898857 |
| | gDNA | 5457 | | | | 7273 | | | | 10992 | | | | 23722 | **23722** | **23722** | |
| Exonic non-synonymous | cfDNA | 4557 | 83 | 0 | 817 | 0 | 38 | 6809 | 426 | 143 | 10335 | 39 | 475 | **303** | **1.2772953** | 1718 | 7.242222 |
| | uDNA | 4142 | 159 | 0 | 1156 | 0 | 167 | 6910 | 196 | 96 | 10509 | 28 | 359 | **450** | **1.8969733** | 1711 | 7.212714 |
| | gDNA | 1320 | | | | 2035 | | | | 3184 | | | | 6539 | **6539** | **6539** | |
| rgeted Genes (5 | cfDNA | 1142 | 24 | 0 | 154 | 0 | 16 | 1930 | 89 | 41 | 2981 | 16 | 146 | **97** | **1.4834072** | 389 | 5.948922 |
| | uDNA | 1006 | 41 | 0 | 273 | 0 | 65 | 1887 | 83 | 19 | 2995 | 8 | 162 | **133** | **2.0339501** | 518 | 7.921701 |
